# A foundation transformer model with self-supervised learning for ECG-based assessment of cardiac and coronary function

**DOI:** 10.1101/2023.10.25.23297552

**Authors:** Jonathan B. Moody, Alexis Poitrasson-Rivière, Jennifer M. Renaud, Tomoe Hagio, Fares Alahdab, Mouaz H. Al-Mallah, Michael D. Vanderver, Sascha N. Goonewardena, Edward P. Ficaro, Venkatesh L. Murthy

## Abstract

**Background:** The wide availability of labeled electrocardiogram (ECG) data has driven major advances in artificial intelligence (AI)-based detection of structural and functional cardiac abnormalities and thus ECG-based diagnosis. However, many critical, high value clinical diagnostic applications, such as assessing myocardial ischemia and coronary microvascular dysfunction, remain underserved due to the limited availability of labeled datasets. We developed a self-supervised ECG foundation model and demonstrate how this approach can overcome this limitation.

**Methods:** A modified vision transformer model was pretrained using a large database of unlabeled ECG waveforms (MIMIC-IV-ECG, N=800,035). The model was then fine-tuned using smaller databases that included high-quality labels derived from positron emission tomography (N=3,126) and clinical reports (N=13,704) for 12 clinical, demographic, and traditional ECG prediction tasks. Diagnostic accuracy and model generalizability were evaluated across five additional cohorts including the publicly available PTB-XL and UK Biobank databases and labels from cardiac magnetic resonance imaging (MRI) and single photon emission computed tomography (SPECT).

**Results:** Diagnostic performance varied across tasks with area under the receiver operating characteristic curve (AUROC) ranging from 0.763 for detection of impaired myocardial flow reserve (MFR < 2) to 0.955 for impaired left ventricular ejection fraction (LVEF < 35%). Self-supervised learning (SSL) pretraining greatly improved diagnostic accuracy in 11 of the 12 prediction tasks compared to conventional *de novo* supervised training. The model retained strong performance across three external and two internal cross-modality databases, with AUROC ranging from 0.771 for impaired MFR to 0.949 for impaired LVEF.

**Conclusion:** This versatile ECG foundation model demonstrates that SSL pretraining enhances diagnostic accuracy and generalizability across diverse cardiac diagnostic applications. By enabling effective learning from limited labeled data, this approach supports AI development for complex but clinically critical tasks, such as detecting myocardial ischemia and coronary microvascular dysfunction, where high-quality labels are costly and scarce.

**Description:** This study introduces a self-supervised ECG transformer foundation model that achieves high diagnostic accuracy across multiple cardiac prediction tasks. By enabling robust performance with limited labeled data, the approach expands AI’s potential in complex diagnostic areas such as myocardial ischemia and coronary microvascular dysfunction.

## Introduction

Nearly 1 in 17 emergency department visits in the United States are for chest pain, representing almost 8 million encounters annually.^1^ Chest pain also drives nearly 6 million outpatient clinic visits per year.^2^ Cardiac stress testing is the dominant diagnostic approach for evaluating such patients.^3^ Several noninvasive testing modalities provide important diagnostic measures of cardiac and coronary function, albeit with varying accessibility and cost (Figure 1). AI models of electrocardiograms (ECGs) have excelled at automating ECG interpretation and show promise as low-cost diagnostic tools.^4^ Because of the wide availability of ground truth annotations from clinical ECG and echocardiographic findings, supervised learning using large, labeled databases has been the dominant paradigm for nearly all of these applications.^5^

**Figure 1:**
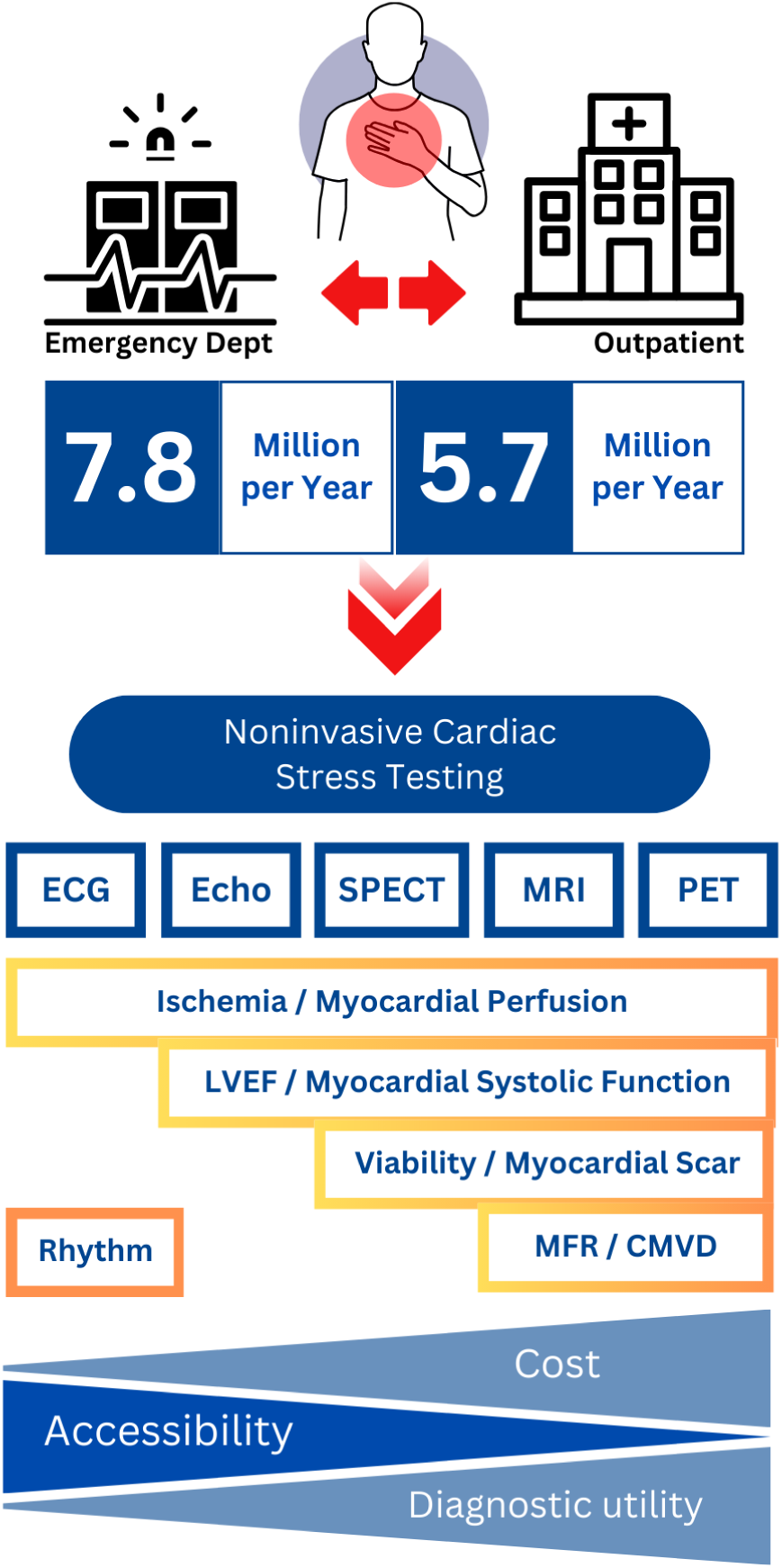
Chest pain encounters in the United States and available noninvasive testing modalities. From left to right, as modality cost increases and accessibility decreases, the traditional capabilities and diagnostic utility for assessing cardiac and coronary function tend to increase.

ECGs can also detect myocardial ischemia and infarction, providing insight into perfusion and potassium balance, which inform the management of cardiovascular disease. Positron emission tomography (PET) myocardial perfusion imaging (MPI) is the gold standard for accurate characterization of ischemia and coronary microvascular dysfunction (CMVD).^6,7^ Such testing enables quantitative cardiac and cardiovascular measures — most importantly, myocardial flow reserve (MFR), which has independent diagnostic and prognostic utility for known or suspected coronary artery disease.^8–10^ However, PET MPI is costly and not widely available,^11^ limiting the availability of data derived from quantitative PET measurements. Consequently, AI-enhanced approaches in this arena have been very limited.

Foundation models and self-supervised learning (SSL) have enabled paradigm shifts across many AI domains.^12^ Foundation models are trained on broad data that can be adapted to a range of downstream tasks.^13^ SSL enables such models to learn powerful task-agnostic representations from large unlabeled databases, allowing for efficient fine-tuning using smaller task-specific annotated databases.^14^ This approach underlies many state-of-the-art large language and computer vision models.^15,16^

Despite these strengths, foundation models and SSL methods have yet to be widely applied to ECG signals. A related AI innovation, the transformer architecture,^17^ now standard in natural language processing,^18^ excels at modeling long-range dependencies in sequential data but has rarely been applied to ECGs.^19,20^

In this work, we developed an SSL foundation model based on a modified vision transformer,^21^ and pretrained it using a generative pretext task in a large database of 800,035 unlabeled ECG waveforms. The model was then fine-tuned in smaller labeled databases for 12 demographic and clinical prediction tasks across 3 major dimensions of cardiology: myocardial function, coronary perfusion, and cardiac rhythm. Four tasks focused on assessing myocardial ischemia and coronary microvascular and vasomotor dysfunction (CMVD), which have been particularly inaccessible with ECG-AI models. We demonstrate the efficacy of the SSL approach by comparing the foundation model with an analogous model developed by conventional *de novo* supervised training. The models were evaluated across three external and two internal cross-modality databases for diagnostic accuracy and generalizability. Results show that combining SSL with a transformer architecture enables efficient training and robust diagnostic performance of high-value, clinically impactful ECG-based AI tools with limited labeled data.

## Methods

### Data sources

Seven ECG databases were used for model development, validation, fine-tuning, and testing Table 1. The first was the publicly available MIMIC-IV-ECG repository v1.0^22,23^ of standard 12-lead resting ECGs of 10 seconds duration. These waveforms were used as unlabeled data for pretraining the SSL foundation model.

**Table 1:**
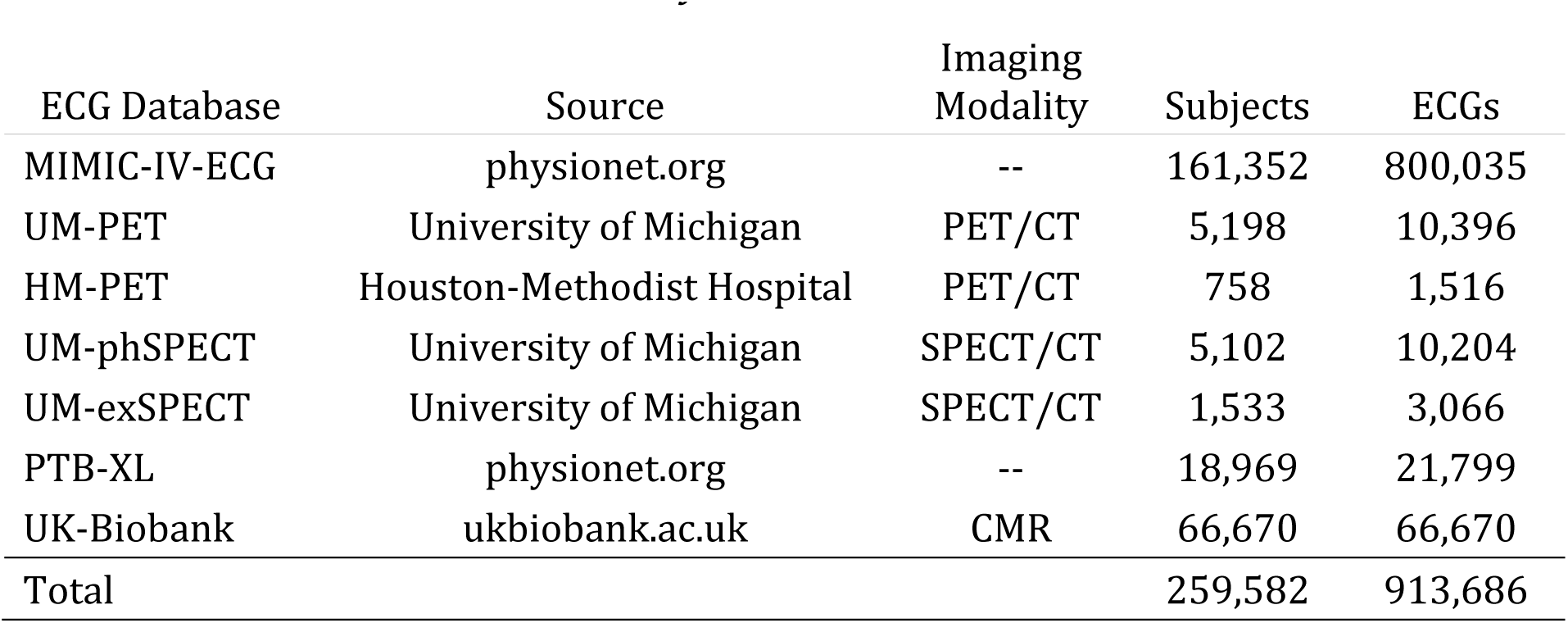
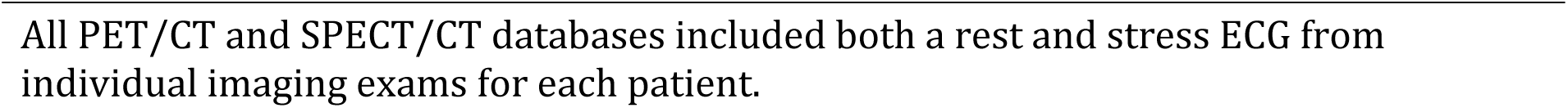
ECG databases used in this study.

| ECG Database | Source | Imaging Modality | Subjects | ECGs |
| --- | --- | --- | --- | --- |
| MIMIC-IV-ECG | physionet.org | -- | 161,352 | 800,035 |
| UM-PET | University of Michigan | PET/CT | 5,198 | 10,396 |
| HM-PET | Houston-Methodist Hospital | PET/CT | 758 | 1,516 |
| UM-phSPECT | University of Michigan | SPECT/CT | 5,102 | 10,204 |
| UM-exSPECT | University of Michigan | SPECT/CT | 1,533 | 3,066 |
| PTB-XL | physionet.org | -- | 18,969 | 21,799 |
| UK-Biobank | ukbiobank.ac.uk | CMR | 66,670 | 66,670 |
| Total |  |  | 259,582 | 913,686 |

All PET/CT and SPECT/CT databases included both a rest and stress ECG from individual imaging exams for each patient.

Three ECG databases were obtained from MPI registries at the University of Michigan Frankel Cardiovascular Center. These consisted of all patients who underwent clinically indicated stress testing with ECG and concurrent noninvasive MPI with either pharmacologic stress positron emission tomography/computed tomography (PET/CT) (UM-PET) or single photon emission computed tomography (SPECT) using pharmacologic (UM-phSPECT) or exercise (UM-exSPECT) stress. The UM-PET database was used for model development, internal validation, and fine-tuning and the two SPECT databases for model evaluation and internal, cross-modality testing.

Quantitative measurements of gold standard PET-MPI data were used to generate ECG labels for task-specific fine-tuning of the foundation model. The PET data were randomly split at the patient level (not study level) into training, validation, and holdout test subsets with the ratios ∼60:20:20% (N=3126/1041/1031). All three Michigan cohorts were reviewed to remove any patient-level overlap. Patients who had undergone both PET/CT and SPECT/CT exams were included only in the UM-PET cohort and excluded from SPECT cohorts to avoid label leakage. Only the earliest evaluable exam was included for patients with more than one MPI exam. Patients were excluded if they had a history of heart transplantation, missing or uninterpretable image data, or missing demographic or hemodynamic data. All patient data was de-identified and informed consent was waived under an exemption from the University of Michigan Institutional Review Board.

The fifth database was a cohort of clinical patients undergoing routine PET-MPI stress testing at the Houston-Methodist hospital and was used for external testing of PET-MPI prediction tasks (HM-PET).

The sixth database was the publicly available PTB-XL ECG database.^24,25^ The predefined folds of this data were used to fine-tune and evaluate the SSL foundation model for automatic ECG interpretation and diagnosis tasks. The entire dataset was also used for externally testing the SSL model fine-tuned on the UM-PET database for the three demographic prediction tasks.

The last database was the publicly available UK Biobank database^26^ which was used for external testing of left ventricular ejection fraction (LVEF) and demographic tasks.

### Noninvasive stress testing

Routine rest and stress cardiac ^82^Rb PET exams were performed in accordance with clinical guidelines for MPI testing^27^ and measurement of myocardial blood flow (MBF)^28,29^ as previously described.^30^

Cardiac ^99m^Tc-sestamibi SPECT exams were performed in accordance with guidelines^31^ as previously described.^32^ LVEF^33^ and stress total perfusion deficit (TPD)^34^ were routinely estimated for all imaging exams. Pharmacologic stress was induced in the UM-PET database with intravenous bolus administration of regadenoson (0.4 mg) and in the UM-phSPECT database with either regadenoson alone or in combination with low-level treadmill exercise. In the UM-exSPECT database, stress was induced by treadmill exercise using a Bruce, modified Bruce, or Cornell protocol. Heart rate, systolic, and diastolic blood pressure were monitored continuously during imaging.

Twelve-lead ECG waveforms of 10 second duration and 500 Hz sampling rate were recorded at baseline immediately before stress testing and at 1-minute intervals during stress (in supine position for pharmacologic or combination pharmacologic/low-level exercise stress) or once during each exercise stage (for exercise stress). Both rest and stress ECG waveforms accompanying each PET MPI exam were used for ECG-AI model development.

### Data annotations

ECG annotations were derived for stress and resting ECGs acquired simultaneously with PET-MPI (nine prediction tasks) or SPECT-MPI (five tasks); for resting ECGs acquired on the same day as cardiac magnetic resonance imaging (MRI) in the UK Biobank data (four tasks); and for resting ECGs in the PTB-XL data (six tasks) Table 2.

**Table 2:** Prediction tasks and ECG databases used for pretraining, fine-tuning and testing the SSL foundation model (P=SSL pretraining, F=Fine-tuning, H=Holdout testing, E=External testing, O=Internal, cross-modality testing).

| Task | MIMIC<br>-IV-ECG | UM-PET | HM-PET | UM-phSPEC<br>T | UM-exSPECT | PTB-XL | UK-Biobank |
| --- | --- | --- | --- | --- | --- | --- | --- |
| **LV Myocardial Perfusion** |  |  |  |  |  |  |  |
| MFR | P | F/H | E | — | — | — | — |
| Rest MBF | P | F/H | E | — | — | — | — |
| Stress MBF | P | F/H | E | — | — | — | — |
| Stress TPD | P | F/H | E | O | O | — | — |
| **LV Myocardial Function** |  |  |  |  |  |  |  |
| LVEF | P | F/H | E | O | O | — | E |
| LVEF Reserve | P | F/H | E | — | — | — | — |
| **Auto ECG Interpretation** |  |  |  |  |  |  |  |
| Diagnostic | P | — | — | — | — | F/H | — |
| Rhythm | P | — | — | — | — | F/H | — |
| Form | P | — | — | — | — | F/H | — |
| **Demographics** |  |  |  |  |  |  |  |
| Age | P | F/H | E | O | O | E | E |
| BMI | P | F/H | E | O | O | E | E |
| Sex | P | F/H | E | O | O | E | E |

MIMIC-IV-ECG data was unlabeled and used for self-supervised pretraining of foundation model.
Labels for LV myocardial perfusion and function tasks were derived from PET imaging.
**MFR**=myocardial flow reserve (threshold < 2).
**Rest MBF**=rest myocardial blood flow (threshold > 1.2 ml/min/g).
**Stress MBF** threshold < 1.5 ml/min/g.
**Stress TPD**=stress total perfusion deficit (threshold > 5%).
**LVEF**=left ventricular ejection fraction (threshold < 35%).
**LVEF Reserve** threshold < -5%.
Auto ECG Interpretation labels derived from clinical ECG reports.

Absolute MBF in ml/min/g of tissue was measured from 30-frame dynamic PET images at rest and stress with a clinically validated, commercially available one-tissue compartmental model, and MFR was computed as the ratio of stress over rest MBF and averaged globally over the entire left ventricular myocardium.^35,36^ Stress TPD was computed from relative myocardial perfusion images as previously described.^37^

Left ventricular volumes were measured from 16-frame ECG-gated myocardial perfusion images using a clinically validated, commercially available algorithm, and LVEF was computed as one minus the left ventricular (LV) volume ratio at systole over diastole.^33^ LVEF reserve was computed as the difference between stress and rest LVEF.^38^ In the UK Biobank database, LVEF measurements derived from cardiac MRI were obtained from the publicly available database.^39,40^

These six tasks were treated as individual single label binary classifications with appropriate thresholds for continuous measures obtained from the literature and clinical experience (MFR < 2, stress MBF < 1.5 ml/min/g, rest MBF > 1.2 ml/min/g,^28^ stress TPD > 5%,^41^ LVEF < 35%,^42^ LVEF reserve < −5%^38^).

Three auto ECG interpretation tasks were treated as a multilabel classification tasks with three categories of ECG statements as defined in the PTB-XL dataset: 44 diagnostic statements, 12 rhythm statements, and 19 form statements.^24^

For the three demographic prediction tasks, age and BMI were treated as continuous regression tasks, and sex as a binary classification task with labels determined by patient records.

### Deep learning model

#### Model development

The primary inputs for the SSL foundation model were raw 12-lead ECG waveforms of 10 second duration. A modified vision transformer neural network^21^ was developed by adapting the initial patch embedding layer for one-dimensional, multichannel input (1dViT) (Supplementary Figure S1 (a)). Waveform patches of fixed 100 millisecond duration corresponding to a patch size of 50 were flattened, linearly projected into an embedding space of dimension 800, combined with a trainable class token, and summed with trainable positional encoding.^21^ A series of 12 transformer encoder blocks followed, each incorporating pre-layer normalization,^43^ 8 self-attention heads, and a multi-layer perceptron with GELU activation^44^ and hidden dimension of 3200 (Supplementary Figure S1 (a)). The final classifier head consisted of global average pooling, layer normalization, and a linear classifier.

The number of input ECG leads was reduced from 12 to 8 (2 bipolar and 6 precordial) since the 3 augmented unipolar leads are linear combinations of the bipolar leads, and by Einthoven’s triangle equation only 2 of the 3 bipolar leads are linearly independent.^45^ A fusion model for dual input using resting and stress ECG waveforms was constructed by concatenating the outputs of two pretrained 1dViT backbones with classifier head removed (Supplementary Figure S1 (b)) and combining with a final linear classifier. This fusion model was fine-tuned or trained from scratch (see Model fine-tuning below) using end-to-end learning using both stress and rest ECG input for each patient. The 1dViT backbone contained 92.7 million trainable parameters and, except for the modified 1D patch embedding and patch size (50), its architecture is similar to a 2D ViT-B/16.^21^ All models were developed using the PyTorch framework (v2.1.1).^46^ Our code is available at https://github.com/4dm-labs/ecgflow and pretrained model checkpoints are available at https://huggingface.co/4dm-labs.

#### ECG preprocessing

Waveforms from the University of Michigan and Houston-Methodist Hospital underwent routine filtering for powerline interference and low-frequency baseline wander^47^ using vendor software on the stress ECG carts. Waveforms from PTB-XL and UK Biobank databases, which had no prior vendor preprocessing, underwent comparable in-house preprocessing using zero phase distortion bandpass (0.01 - 150 Hz)^48^ and notch filtering (0.001 Hz). A small number of waveforms from the MIMIC-IV-ECG databases had missing sample points which were imputed using the mean. Each waveform lead was additionally normalized to zero mean and unit standard deviation (SD) using the grand mean and SD of either the MIMIC-IV-ECG database during SSL pretraining or the UM-PET ECG training database during fine-tuning. The grand mean (SD) in the MIMIC-IV-ECG training split (N=780,035) for 8 leads was: 0.0099 (0.2166) mV. This was similar to the grand mean (SD) in the UM-PET training split (also in physical mV units): 0.0153 (0.2333).

#### SSL model pretraining

We adapted for 1D ECG data a particularly simple generative SSL methodology called the masked auto-encoder.^14,49^ For SSL pretraining, the 1dViT backbone without the classifier head was paired with a small transformer decoder to perform a pretext task of masked signal modeling.^50,51^ The decoder had eight transformer blocks similar to the 1dViT encoder but with embedding dimension of 256 and 16 self-attention heads.^51^ Embedded input patches were randomly masked with a masking ratio of 60%, and only unmasked patches were input to the encoder.^51^ Before entering to the decoder, encoded patches were combined with a trainable token for each masked patch and summed with positional encoding. The input waveform was reconstructed at the level of raw time steps with an *L*_2_ loss.^51^ Our pretraining method closely follows generative SSL methods from the literature.^21,50,51^

The model was trained using the AdamW optimizer^52^ with an initial learning rate of 1e-5, linear warmup for five epochs to a peak of 1e-3 followed by a cosine decay schedule.^53^ Pretraining was performed for 300 epochs with a batch size of 384.

#### Model fine-tuning

The SSL pretrained 1dViT encoder was subsequently fine-tuned on labeled data for each prediction task. In addition to the fusion 1dViT for dual stress-rest ECG input, a single 1dViT model was also fine-tuned for rest-only ECG input to evaluate the additional performance provided by using stress ECGs. Only the classifier head and top four transformer blocks were trained during fine-tuning, while freezing the remaining eight encoder blocks. A binary cross-entropy loss function was utilized for multi- and single-label tasks and mean squared error loss for continuous regression tasks. We used the AdamW optimizer with linear warmup of the learning rate from 1e-5 to a peak of 5e-5 followed by cosine decay. Model and training hyperparameters were set using suggested values from the literature.^21,54^ Label smoothing regularization^55^ was tuned to optimize model calibration^56^ by minimizing the mean absolute difference and 90^th^ percentile absolute difference between observed and predicted probabilities — integrated calibration index (ICI) and E90.^57^ For the auto ECG interpretation tasks, positive label weights were used to compensate for label imbalance during training and fine-tuning. Fine-tuning was performed to maximize area under the receiver operating characteristic curve (AUROC) using 100 epochs with a batch size of 256. Training iterations were stopped early when validation AUROC failed to improve for ten consecutive epochs. For comparison, the same model without SSL-pretraining was trained and validated *de novo* from random initialization with the same labeled data and training setup, except the peak learning rate during linear warmup was set to 1e-3 as used for SSL pretraining.^54^ In the same way, fine-tuning of the SSL pretrained model and supervised *de novo* training for the auto ECG interpretation tasks was performed using the PTB-XL predefined training folds.^24^

#### Model evaluation

Feature maps generated at the last transformer block were isolated and projected into two dimensions for cluster analysis using the LocalMAP refinement of the PaCMAP dimension reduction algorithm^58,59^ (Supplementary Figures S14 – S17).

Model discrimination of binary classification tasks was assessed using ROC curves, diagnostic accuracy, AUROC, and macro averages for multi-label tasks. Model calibration for these tasks was assessed graphically with calibration curves of predicted and observed probabilities, and quantitatively using ICI and E90. Continuous regression tasks were evaluated with scatter and Bland-Altman plots of true and predicted values, mean absolute error (MAE), and R^2^. All model evaluations in the UK Biobank and PTB-XL databases used the rest ECG single input 1dViT model since appropriate stress ECGs were not available in these databases (limited 4-lead exercise ECGs that are available in UK Biobank were not adapted here).

Label efficiency was evaluated for five tasks over two databases: LVEF and MFR (UM-PET), and the three auto ECG interpretation tasks (PTB-XL). The pretrained SSL model was fine-tuned over a series of randomly sampled data subsets of decreasing size. For comparison, the same model was also trained *de novo* from random initialization using the same hyperparameter settings described above. Each model was then evaluated with holdout data and the entire process was repeated five times with different random seeds.

### Statistical methods

For diagnostic evaluation, the DeLong test^60^ was used to compare single-label AUROCs, McNemar test^61^ for comparison of sensitivity and specificity, and Leisenring test^62^ for comparison of positive and negative predictive values. Bootstrap methods were used to compare multilabel models.^63^ To facilitate comparison between ROC curves, a threshold was selected at the point on the curve that minimized the distance to the point of perfect discrimination (sensitivity=1, specificity=1). Subgroup analysis of model performance was performed, stratifying by age (< 60 y), BMI (< 30 kg/m^2^), sex, diabetes, dyslipidemia, hypertension, known coronary artery disease (CAD), LVEF (< 50%), and stress TPD (< 5%).

Continuous variables are summarized as mean±SD or median [1st – 3rd quartiles]. Welch’s unequal variances t-test or Wilcoxon rank sum tests were used as appropriate for comparisons of continuous parameters, and *χ*-squared tests were used to compare categorical variables. Two-sided p-values less than 0.05 were considered statistically significant.

## Results

### Patient populations

The MIMIC-IV-ECG repository provided 800,035 diagnostic resting ECGs acquired in 161,352 patients. For SSL pretraining, ECG data were split into training/validation subsets 780,035/20,000 (97.5%/2.5%) and no other patient data were used. In the UM-PET database, after excluding 259 heart transplant patients and 4 PET/CT patients due to uninterpretable PET data or missing demographic data, 5,198 patients were available for ECG-AI model development and testing: 3,126 (training or fine-tuning), 1,041 (validation), and 1,031 (holdout test) (Supplementary Figure S2). The sizes of evaluation databases are listed in Table 1.

### Baseline characteristics

Baseline characteristics are shown in Table 3 stratified by patient cohort. As expected, the PET MPI cohorts for model derivation and holdout test had nearly identical demographics and risk factor profiles. However, the pharmacologic stress SPECT MPI testing cohort differed meaningfully from PET MPI cohorts. SPECT patients were older, with lower rates of obesity, diabetes, and history of MI, and higher rates of hypertension, hyperlipidemia, and mortality. Baseline characteristics of the exercise stress SPECT MPI cohort are compared in Supplementary Table S1. The number of positive cases for each task and cohort (overall and within subgroups) is shown in Supplementary Figures S6 - S10. In particular, the prevalence of impaired LVEF was very low in the UM-exSPECT (0.5%) and UK Biobank (0.3%) cohorts.

**Table 3:** Baseline characteristics of patients who underwent stress testing by myocardial perfusion imaging in the UM-PET and UM-phSPECT databases.

|  | UM-PET |  | UM-phSPECT | p-value |
| --- | --- | --- | --- | --- |
|  | Model Derivation | Holdout Test | Internal Test |  |
| n | 4167 | 1031 | 5102 |  |
| Age (y) | 63 (12) | 63 (12) | 66 (12) | <0.001 |
| Male Sex | 2312 (55) | 590 (57) | 2874 (56) | <0.001 |
| White | 3404 (82) | 840 (81) | 4127 (81) | <0.001 |
| Black | 512 (12) | 115 (11) | 504 (10) |  |
| Other | 251 (6) | 76 (7) | 471 (9) |  |
| Body mass index (kg/m<sup>2</sup>) | 34 (10) | 33 (9) | 30 (6) | <0.001 |
| Obesity | 2539 (61) | 599 (58) | 2303 (45) | <0.001 |
| Hypertension | 3179 (76) | 776 (75) | 4017 (79) | <0.001 |
| Diabetes | 1652 (40) | 422 (41) | 1676 (33) | <0.001 |
| Hyperlipidemia | 2747 (66) | 673 (65) | 3699 (73) | <0.001 |
| Family history of coronary artery disease | 2807 (67) | 681 (66) | 1715 (34) | <0.001 |
| Current smoker | 336 (8) | 99 (10) | 1135 (22) | <0.001 |
| History of myocardial infarction | 799 (19) | 200 (19) | 714 (14) | <0.001 |
| Prior percutaneous coronary intervention | 816 (20) | 201 (19) | 952 (19) | <0.001 |
| Prior coronary artery bypass graft | 418 (10) | 102 (10) | 491 (10) | <0.001 |
| Angina | 1819 (44) | 431 (42) | 2204 (43) | <0.001 |
| Dyspnea | 665 (16) | 172 (17) | 1077 (21) | <0.001 |
| Heart rate, rest (bpm) | 73 (14) | 73 (13) | 69 (13) | <0.001 |
| SBP, rest (mm Hg) | 132 (23) | 131 (24) | 143 (25) | <0.001 |
| DBP, rest (mm Hg) | 71 (24) | 71 (22) | 80 (13) | <0.001 |
| Rate pressure product (mm Hg bpm) | 9658 (2496) | 9527 (2470) | 9853 (2522) | <0.001 |
| Heart rate, stress (bpm) | 93 (20) | 92 (16) | 101 (20) | <0.001 |
| SBP, stress (mm Hg) | 120 (31) | 119 (30) | 161 (27) | <0.001 |
| DBP, stress (mm Hg) | 64 (39) | 63 (36) | 80 (14) | <0.001 |
| LV ejection fraction,<br>rest (%) | 57 (15) | 58 (15) | — | 0.806 |
| LV ejection fraction,<br>stress (%) | 61 (16) | 61 (16) | 65 (13) | <0.001 |
| Total perfusion deficit,<br>stress (%) | 7 (10) | 7 (11) | 5 (9) | <0.001 |
| Myocardial blood flow,<br>rest (ml/min/g) | 1.08 (0.39) | 1.08 (0.41) | — | 0.929 |
| Myocardial blood flow,<br>stress (ml/min/g) | 2.17 (0.83) | 2.16 (0.82) | — | 0.784 |
| Myocardial flow reserve | 2.11 (0.72) | 2.12 (0.74) | — | 0.884 |

### SSL pretraining

Figure 2 compares ECG-AI model performance with and without SSL pretraining for nine tasks evaluated in the external HM-PET database. SSL pretraining produced significant AUROC increases from +0.055 to +0.103 for all binary classification tasks except prediction of impaired LVEF reserve (+0.006, *p* = 0.850; see Supplementary Tables S2, S3, and S4 for complete summaries including point estimates and 95% confidence intervals for each evaluation metric). In addition, R^2^ was increased and MAE reduced for continuous regression tasks (Supplementary Table S2). Additional testing of these same tasks evaluated in the UM-PET holdout database yielded similar diagnostic performance and SSL benefits (Supplementary Figure S3, Supplementary Table S4). Strong diagnostic performance was maintained in internal, cross-modality tests evaluated in the pharmacologic (UM-phSPECT) and exercise (UM-exSPECT) stress SPECT cohorts, and external tests in the UK Biobank and PTB-XL databases (Figure 3) (labels were not available for every task).

**Figure 2:**
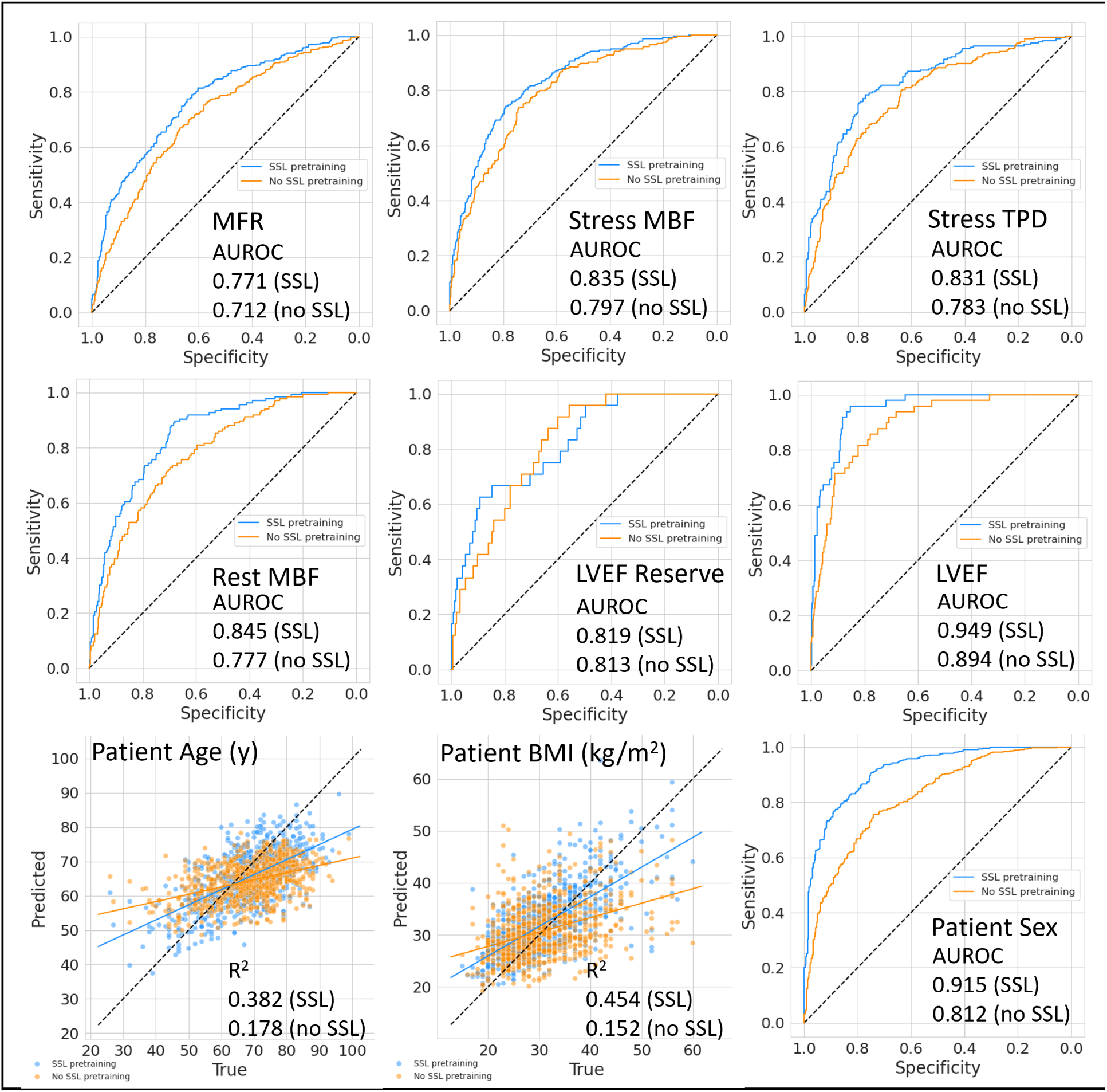
External tests evaluated in the HM-PET database. Task performance was compared with and without SSL pretraining. The SSL pretrained model was fine-tuned in the UM-PET model derivation cohort. The non-SSL model underwent supervised de novo training from random initialization in the same UM-PET cohort. A complete summary is given in Supplementary Table S2.

**Figure 3:**
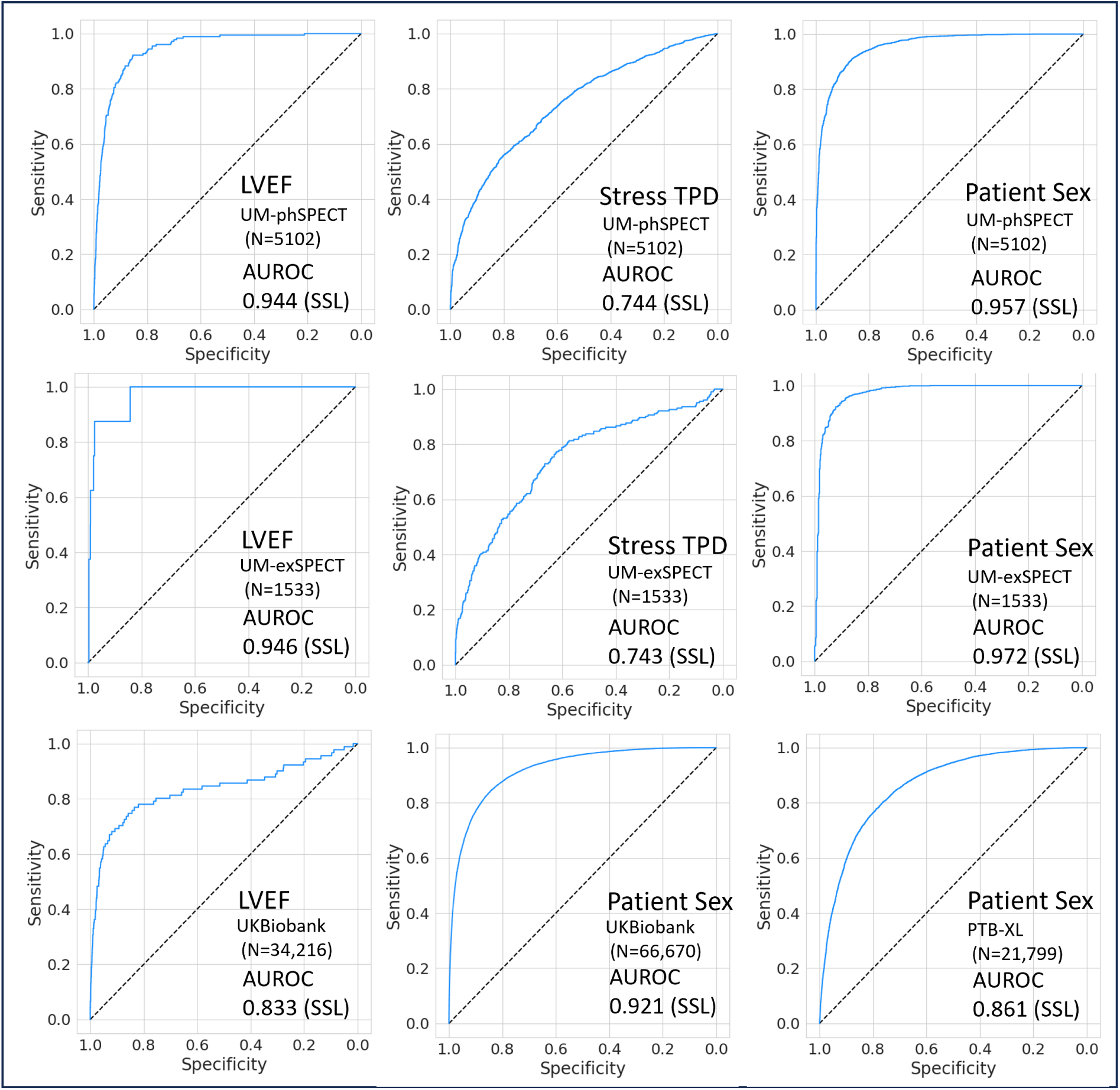
Additional tests of the SSL pretrained model in the UM-phSPECT, UM-exSPECT, UK Biobank, and PTB-XL databases.

Supplementary Table S3 and Supplementary Figure S4 compare ECG-AI model performance for three auto ECG interpretation tasks in the PTB-XL database. The increased performance with SSL pretraining was maintained in terms of macro AUROC across all tasks (+0.035 to +0.129, *p* < 0.0097), as well as macro Youden index for two of the three tasks (+0.100 to +0.219, *p* < 0.0089).

Label efficiency of the SSL-pretrained model is compared to *de novo* training in Figure 4 for four tasks across two databases. Label efficiency varied by task and database but generally improved with SSL pretraining. The *de novo* model AUROC tended to plateau at < 100% of the training data while the SSL-pretrained model continued to increase. In all four tasks the SSL model reached the highest AUROC achieved by the *de novo* model with far less than 100% of the training data.

**Figure 4:**
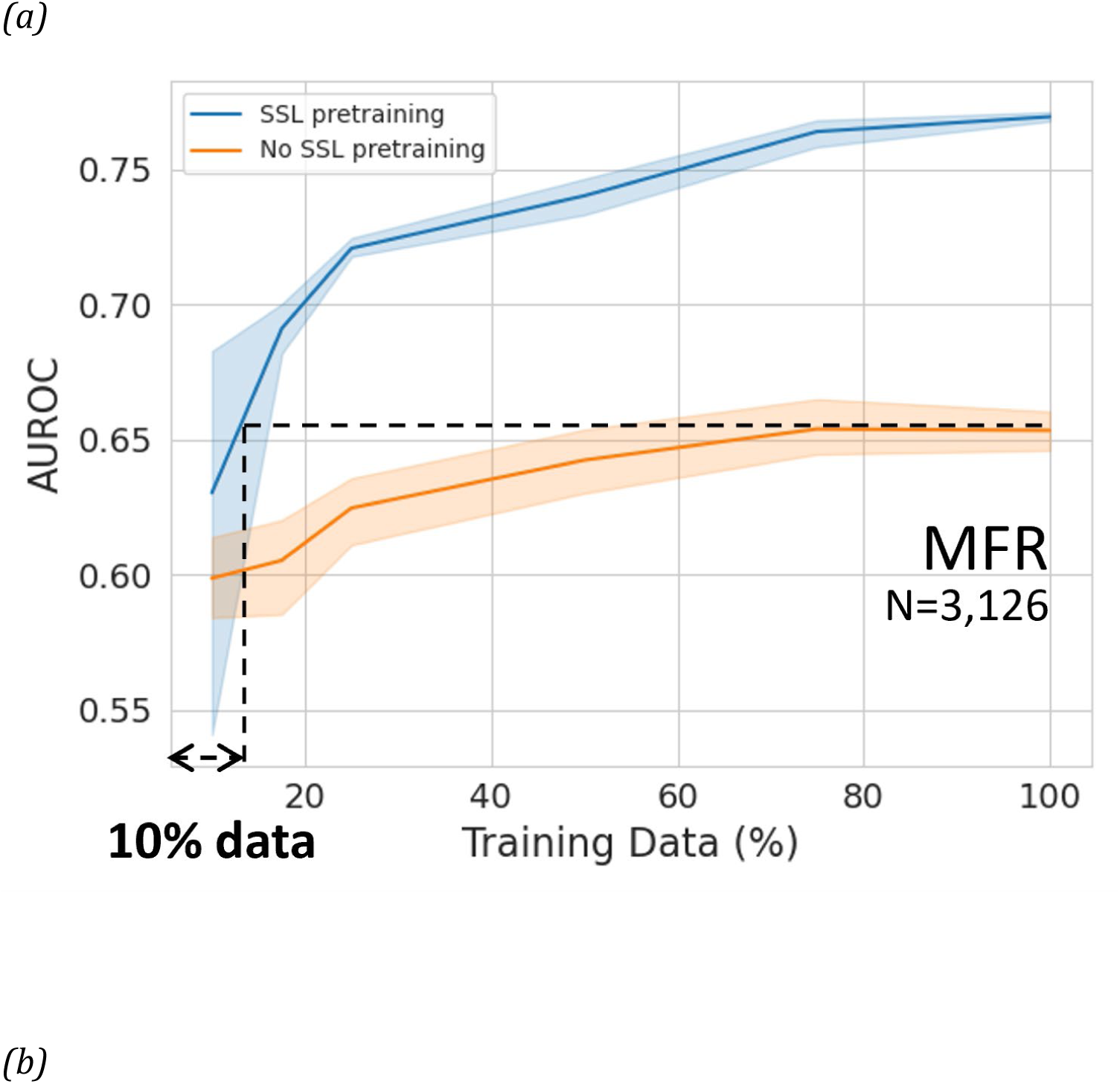
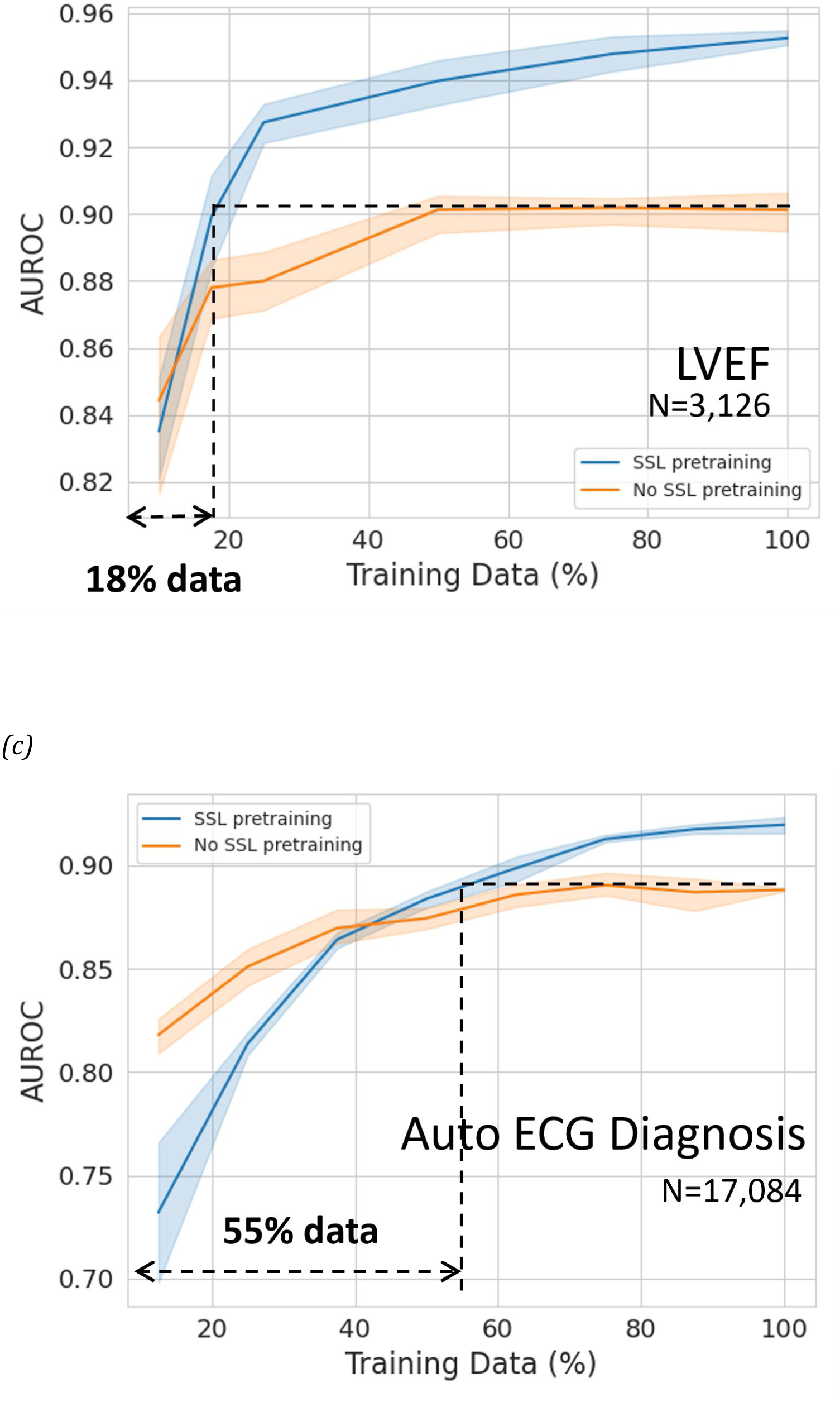
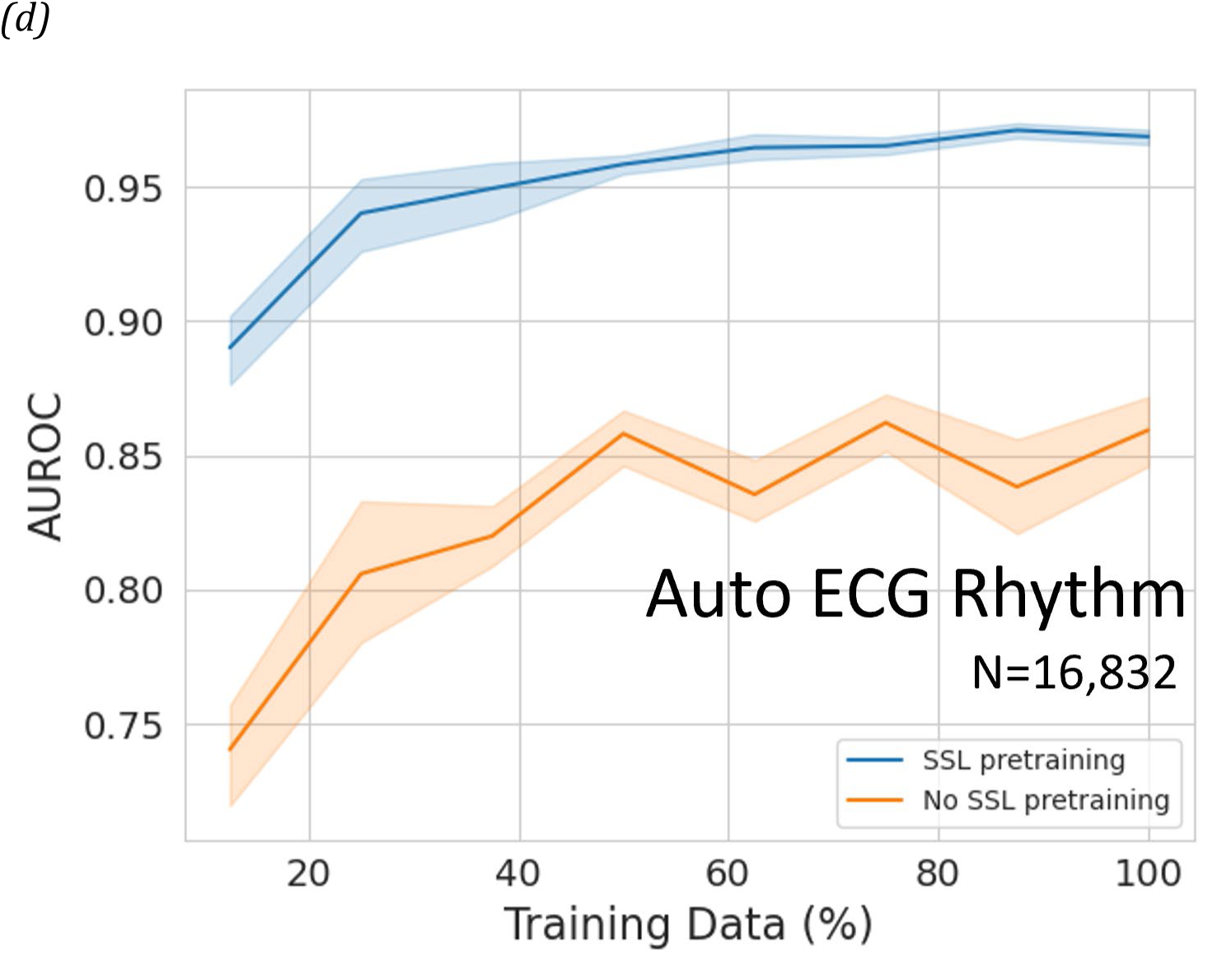
Evaluation of label efficiency. AUROC versus the fraction of training data used for supervised fine-tuning after SSL pretraining and de novo training with no SSL pretraining. (a) MFR task and (b) LVEF task in the UM-PET holdout database; (c) auto ECG diagnosis task and (d) auto ECG rhythm task in the PTB-XL database.

Two examples of model calibration are shown in Supplementary Figure S5. Model calibration was improved in terms of ICI and E90 after tuning label smoothing regularization.

Subgroup analysis demonstrated broadly consistent model performance over multiple tasks and evaluation databases (Supplementary Figure S6 to Supplementary Figure S10).

### Benefits of stress ECG

Some performance increases were observed for the dual input (stress-rest ECG) 1dViT model compared to the single-input (rest-only ECT) model. However, they tended to be more modest in magnitude and scope than that of SSL pretraining. AUROC improved for the stress MBF task (rest-only ECG 0.841 95% CI [0.812 - 0.870] vs. stress-rest ECG 0.866 95% CI [0.840 - 0.892], *p* = 0.0052) and stress TPD (rest-only ECG 0.740 [0.707 – 0.772] vs. stress-rest ECG 0.762 [0.730 – 0.794], *p* = 0.0411). However, changes in sensitivity and specificity using stress-rest ECGs were mixed (sensitivity: MFR +0.061 [0.0256 – 0.0956]; stress MBF +0.005 [-0.0371 – 0.0464]; LVEF +0.062 [0.0056 – 0.1183]; specificity: MFR - 0.047 [-0.0754 – −0.0179]; stress MBF +0.056 [0.0334 – 0.0793]; LVEF −0.053 [-0.0697 – - 0.0371]). Note that all external evaluations with UK Biobank and PTB-XL databases (including SSL vs. non-SSL) used the single-input rest-only model as stress ECGs were not available.

## Discussion

We applied a label efficient SSL approach to implement a foundation model capable of performing multiple clinically relevant ECG-based prediction tasks with limited labeled data. The model was pretrained with a self-supervised learning method on a large publicly available database of unlabeled ECG data and was then fine-tuned on smaller datasets with labels derived from PET, patient demographics, and clinical ECG reports targeting 12 prediction tasks. The model was validated in five additional datasets that included labels derived from two additional imaging modalities (MRI and SPECT). Compared with *de novo* supervised training, the SSL paradigm consistently improved the diagnostic accuracy of 11 of the 12 prediction tasks investigated. Importantly, this approach may facilitate the development of deep learning-based, low-cost testing strategies for more complex cardiac diagnostics previously thought to be inaccessible due to the need for extremely large training datasets.

### ECG-AI prediction tasks

Out of our 12 ECG prediction tasks, 7 have been previously studied. ECG-AI prediction of LVEF was reported by Attia et al.^42^ and has been externally validated in multiple prospective cohorts.^64^ In our small, PET-derived LVEF dataset, we observed remarkably high diagnostic accuracy after *de novo* supervised training, which was significantly improved with SSL pretraining and fine-tuning, and remained consistently high in external and internal cross-modality testing.

ECG-AI prediction of age, BMI, and sex has been reported by several groups,^63,65–68^ under the hypothesis that differences between predicted and actual values reflect general cardiovascular or cardiometabolic health, or hormonal status.^69^ Again, we observed quantitative and diagnostic accuracy comparable to prior studies,^63,67,68^ including in two publicly available datasets (PTB-XL and UK Biobank), and the accuracy was further improved by SSL pretraining. Similarly, the SSL foundation model fine-tuned in the PTB-XL database also increased diagnostic accuracy for three auto ECG interpretation tasks compared with *de novo* training.

Five of our prediction tasks leveraged rest and stress PET-derived labels that have not been previously studied with deep neural network ECG modeling. Stress TPD is a relative measure of myocardial perfusion that indicates the extent and severity of focal obstructive epicardial disease.^34^ Stress and rest MBF and MFR (MFR = stress MBF divided by rest MBF) are measures that provide complimentary information and add independent, incremental diagnostic and prognostic value to stress TPD measurements.^70,71^ LVEF reserve provides additional information on the presence of high-risk multi-vessel disease.^38,72^ Together these PET-derived measures provide comprehensive, state-of-the-art assessment of left ventricular myocardial and coronary function in patients with known or suspected coronary artery disease. Additionally, since stress and rest ECGs were acquired simultaneously with each PET scan, these measures provide high-quality data annotations for ECG model training and fine-tuning.

In addition to external testing of the ECG-AI model with cardiac PET data from the Houston-Methodist Hospital, further testing was performed in four other databases (UK Biobank, PTB-XL, UM-phSPECT, and UM-exSPECT). Although the UM-phSPECT and UM-exSPECT databases were collected at the same institution as the UM-PET model derivation database, these cohorts actually represent out-of-distribution datasets with respect to the UM-PET cohort. There were significant differences between these cohorts (Table 3 and Supplementary Table S1) in clinical referral to stress testing; imaging and stress modality; patient demographics, symptoms, and cardiovascular risk factors; resting and stress hemodynamics during image and ECG acquisition; LVEF and incidence of impaired LVEF (< 35%); stress TPD and incidence of abnormal stress TPD (< 5%); and clinical outcomes (Supplementary Figure S12). The 1dViT feature maps also exhibited distinct global structures among these cohorts (Supplementary Figures S16 – S17). Labels were not available for all tasks due to the retrospective nature of the data, and labels for the LVEF and stress TPD tasks were derived from modalities (MRI and SPECT) with significantly different image resolution, noise properties, and contrast compared to the model derivation database (PET). Nevertheless, the SSL pretrained model retained strong diagnostic performance similar to that of UM-PET holdout testing.

### SSL pretraining

A number of different SSL pretraining approaches have been described in the computer vision literature which may be broadly classified as contrastive or generative.^73^ Although these methods have often performed similarly in the image recognition literature, generative approaches have been reported to perform better in the context of multichannel time-series modeling^49^ and 2D retinal image modeling.^14^ Our generative approach is analogous to that of Zhou et al.^14^ which similarly showed strong performance over several prediction tasks. The regularity of these types of data across patients may make contrastive methods less suitable for pretraining, although further investigation is warranted.

### Impact of stress ECG

Four of our tasks address prediction of ischemia-related parameters (MFR, stress MBF, stress TPD, and LVEF reserve) that involve pharmacologic or exercise stress testing and provided both resting and stress ECGs. Although we expect stress perturbations to add important additional information to model predictions for these tasks, we only observed a modest increase in task performance compared to rest-only ECG input. In this retrospective observational study, we were unable to definitively identify a mechanism for the relatively high informational content of resting ECGs compared to stress. Stress ECGs are often subject to more noise and artifacts than resting ECGs and routinely undergo more aggressive filtering which may suppress some useful signals. Further, our SSL pretraining only included resting ECGs from the MIMIC-IV-ECG database. We aim to explore this question in future studies by including stress ECGs in SSL pretraining and further analysis of feature map clustering.

### Related work

Very few ECG-AI models have been reported that address angina patients undergoing stress testing. To our knowledge, our ECG foundation model is the first deep neural network that uses both rest and stress ECGs to detect multiple aspects of impaired myocardial perfusion, confirming that tissue- and molecular-level pathophysiology results in characteristic electrophysiologic changes detectable in surface ECG tracings. Ahmad and colleagues^74^ performed a ground-breaking study on a similar question in patients with suspected CMVD. Using a range of machine learning approaches with resting ECG data, they built prediction models to detect impaired coronary flow reserve, a measure analogous to MFR and identified by invasive coronary vasomotor reactivity testing. Although such invasive testing is precise, it is not routinely performed and can be subject to substantial referral bias. Because of the limited sample size and lack of stress perturbation data, the accuracy of their prediction models was only moderate.^74^ Alahdab et al.^75^ recently reported an ECG machine learning model based on a PET-derived labels to predict MBF and MFR from resting ECGs. In an independent cohort of patients who underwent SPECT MPI, they demonstrated that ML-predicted MFR was significantly associated with risk of major adverse cardiac events. Our results extend these earlier findings and confirm the utility of ECGs for AI-enhanced assessment of myocardial ischemia and CMVD.

Several previous studies have reported high-performing foundation models based on contrastive, token-based SSL-pretraining or conventional supervised learning.^76–78^ These have primarily targeted traditional ECG interpretation, LVEF, and cardiac biomarker tasks and have benefited from very large, private, labeled ECG databases. By contrast, our choice of tasks was largely driven by the desire to leverage high-value cardiac and coronary diagnostics using much smaller, less accessible labeled databases. Vaid et al.^20^ recently reported a foundation model based on the BEiT transformer framework which also uses a token-based self-supervised pretraining method with unlabeled data. However, the BEiT framework operates on 2D images of ECGs rather than the raw waveform signals as used in our model. In 2D benchmark comparisons reported by He et al.^51^ our SSL pretraining method performed somewhat better on downstream tasks than that used by BEiT.

### Clinical implications

Many successful ECG-based AI prediction models have been reported for such tasks as automatic ECG interpretation,^4,5,63,79^ detection of left ventricular (LV) and right ventricular (RV) systolic dysfunction,^42,64,80^ and demographic inference.^68,69^ These studies have been enabled by the accessibility of very large retrospective datasets derived from general ECG or echocardiography populations, and the models are robust in diverse external populations as observed in our study.

In contrast, the smaller stress testing populations considered in this study are underrepresented in the ECG-AI literature, reflecting the difficulties of training strong AI models that address important diagnostic challenges for these patients. These smaller populations are characterized by a high incidence of obesity, diabetes, and other cardiovascular risk factors and comorbidities which are strongly associated with CMVD development.^7^ CMVD causes myocardial ischemia and symptomatic angina without obstructive coronary disease^81,82^ and is associated with markedly increased rates of adverse cardiac outcomes.^10,83^ Despite high prevalence and prognostic relevance,^10^ CMVD is difficult to diagnose with traditional clinical tools, and many patients remain undiagnosed or have presumptive diagnoses without confirmation. Advanced diagnostic measures that can identify CMVD include quantitative coronary or myocardial flow reserve as measured by invasive vasomotor reactivity testing or noninvasive stress imaging with quantitative PET MPI,^9^ transthoracic doppler echocardiography, or cardiovascular MRI.^84^

These advanced techniques are not widely available and are substantially more costly than standard cardiovascular testing methods, such as invasive coronary angiography and exercise stress ECG, which cannot quantify MFR. Among the advanced tests, quantitative PET MPI is considered the noninvasive gold standard for assessing CMVD,^85^ and likely provides the highest quality data labels for AI model training. Although guidelines broadly recommend PET testing for a range of serious clinical conditions,^88^ its accessibility remains limited.^11^ In resource-constrained environments, ECG-AI predicted MFR could provide a cost-effective initial test to direct patients who would benefit most from quantitative PET MPI testing. Further, ECG-AI MFR could be useful as an adjunct to other more accessible stress testing modalities such as stress SPECT MPI, MRI, stress echocardiography, and exercise ECG, providing valuable additional information on CMVD phenotyping.

### Limitations

The present study did not evaluate the diagnostic performance of ECG-AI models for detection of obstructive CAD. Although inaccurate assessment of “balanced” multivessel CAD is a long-standing limitation of standard SPECT MPI,^89^ PET MPI enables much more accurate detection.^90,91^ We speculate that ECG-AI MFR impairment could potentially improve the diagnostic accuracy of MPI in the presence of high-risk left main or multi-vessel obstructive CAD, as has been shown for quantitative PET MFR in prior studies.^92–94^

Global left ventricular PET MFR does not distinguish CMVD from myocardial perfusion impairments due to focal epicardial or diffuse CAD. Our recent work has shown that regional quantitative PET measures combined with relative perfusion and global MFR can be highly effective at assessing the additive risk of diffuse or microvascular disease.^94,95^ Further work will be necessary to integrate these results with the present ECG-AI model predictions to further characterize CMVD.

Although we provide benchmark comparisons with a variety of models from the literature (Supplementary Tables S5-S9), in this study SSL benchmarking with other architectures was not performed.

### Conclusion

Our ECG foundation model using SSL and fine-tuning for multiple clinical tasks with limited labeled data demonstrated strong diagnostic performance in five independent databases.

The model is uniquely capable of identifying impaired MFR, a hallmark of CMVD, demonstrating that a pretrained foundation model can enable AI applications that are currently underdeveloped due to costly and scarce high-quality labeled data.

## Data Availability

Individual subject level data from the University of Michigan and Houston-Methodist Hospital cannot be shared publicly due to medical data privacy regulations.
Other data are available from public repositories as indicated herein.

https://physionet.org/content/mimic-iv-ecg/1.0/

https://www.ukbiobank.ac.uk/

## Acknowledgements

The authors acknowledge the Regents of the University of Michigan for the use of de-identified clinical data for this study. This research has been conducted using the UK Biobank Resource under application number 234540.

## Funding

SNG is supported by VA MERIT grant 1I01CX002560, NIH/NHLBI R01HL150392, and the Taubman Medical Research Institute (Wolfe Scholarship). VLM is supported by grants R01AG059729 from the National Institute on Aging, U01DK123013 from the National Institute of Diabetes and Digestive and Kidney Disease, and R01HL136685 from the National Heart, Lung, and Blood Institute as well as the Melvyn Rubenfire Professorship in Preventive Cardiology.

## Author Declarations

JBM, APR, JMR, TH, and MDV are employees of INVIA. JMR is a consultant for Jubilant Radiopharma and receives royalties from the licensing of FlowQuant software. EPF is a stockholder in INVIA. VLM has received research grants and consulting fees from Siemens Healthineers and serves as a scientific advisor for Ionetix and owns stock options in the same. He owns stock in GE and Cardinal Health and has received payments for consulting from INVIA. FA, MHA, and SNG have nothing to declare.

## Ethics Approval

This is an observational study, and informed consent was waived under an exemption from the University of Michigan Institutional Review Board.

## Notes

### Author Declarations

This is an observational study and informed consent was waived under an exemption from the University of Michigan Institutional Review Board. This research has been conducted using the UK Biobank Resource under application number 234540.

### Summary of Updates

Added data and results on 10 additional prediction tasks. This version has been accepted for publication by NEJM-AI.

