## Supplemental Data for "A foundation transformer model with self-supervised learning for ECG-based assessment of cardiac and coronary function"

### Statistical methods

Analyses were performed using R Statistical Software (v4.4.2)[1] with packages `pROC` (v1.18.0)[2], `DTComPair` (v1.2.4)[3], `gtsummary` (v1.5.2)[4], and `Publish` (v2021.05.25)[5].

### Supplemental Tables

*Supplementary Table S1: Baseline characteristics of patients from two internal cross-modality databases who underwent SPECT myocardial perfusion imaging stratified by stress modality.*

| | Internal Test | | |
|---|---|---|---|
| | UM-phSPECT (Pharma stress) | UM-exSPECT (Exercise stress) | p-value |
| n | 5102 | 1533 | |
| **Demographics** | | | |
| Age (y) | 66 (12) | 61 (11) | <0.001 |
| Male Sex | 2874 (56) | 974 (64) | <0.001 |
| **Race** | | | |
| White | 4127 (81) | 1279 (83) | <0.001 |
| Black | 504 (10) | 89 (6) | |
| Other | 471 (9) | 165 (11) | |
| **Cardiovascular Risk Factors** | | | |
| Body mass index (kg/m$^2$) | 30 (8) | 29 (5) | <0.001 |
| Obesity | 2299 (45) | 593 (39) | <0.001 |
| Hypertension | 4017 (79) | 907 (59) | <0.001 |
| Diabetes | 1676 (33) | 317 (21) | <0.001 |
| Hyperlipidemia | 3699 (73) | 1029 (67) | <0.001 |
| Family history of coronary artery disease | 1715 (34) | 576 (38) | 0.005 |
| Current smoker | 1135 (22) | 207 (14) | <0.001 |
| History of myocardial infarction | 714 (14) | 137 (9) | <0.001 |
| Prior percutaneous coronary intervention | 952 (19) | 242 (16) | 0.011 |
| Prior coronary artery bypass graft | 491 (10) | 68 (4) | <0.001 |
| **Symptoms** | | | |
| Angina | 2204 (43) | 855 (56) | <0.001 |
| Dyspnea | 1077 (21) | 305 (20) | 0.322 |
| **Hemodynamics** | | | |
| Heart rate, rest (bpm) | 69 (13) | 66 (11) | <0.001 |
| SBP, rest (mm Hg) | 143 (25) | 135 (18) | <0.001 |

| | | | |
|---|---|---|---|
| DBP, rest (mm Hg) | 80 (13) | 80 (11) | 0.387 |
| Rate pressure product (mm Hg bpm) | 9853 (2522) | 8920 (2059) | <0.001 |
| Heart rate, stress (bpm) | 101 (21) | 145 (21) | <0.001 |
| SBP, stress (mm Hg) | 161 (27) | 190 (26) | <0.001 |
| DBP, stress (mm Hg) | 80 (14) | 80 (14) | 0.178 |
| **MPI Measurements** | | | |
| LV ejection fraction, stress (%) | 65 (13) | 68 (10) | <0.001 |
| Total perfusion deficit, stress (%) | 5 (9) | 3 (6) | <0.001 |

*Supplementary Table S2: External evaluation in the HM-PET database (N=758 patients). Task performance was compared with and without SSL pretraining. The model with SSL was pretrained using the MIMIC-IV ECG database and underwent supervised fine-tuning in the UM-PET training split. The model without SSL pretraining underwent supervised de novo training from random initialization in the same UM-PET training split.*

| | No SSL Pretraining | | SSL Pretraining | | |
|---|---|---|---|---|---|
| | Estimate | 95% CI | Estimate | 95% CI | p-value |
| **MFR** | | | | | |
| AUROC | 0.712 | (0.675, 0.748) | 0.771 | (0.738, 0.804) | 0.0003 |
| Sensitivity | 0.669 | (0.622, 0.717) | 0.747 | (0.703, 0.791) | 0.0017 |
| Specificity | 0.671 | (0.624, 0.718) | 0.663 | (0.616, 0.710) | 0.7532 |
| PPV | 0.662 | (0.614, 0.710) | 0.681 | (0.636, 0.727) | 0.2913 |
| NPV | 0.678 | (0.631, 0.725) | 0.731 | (0.685, 0.778) | 0.0054 |
| AUPRC | 0.690 | (0.630, 0.743) | 0.761 | (0.711, 0.808) | 0.0004 |
| **Rest MBF** | | | | | |
| AUROC | 0.777 | (0.736, 0.818) | 0.845 | (0.812, 0.879) | <0.0001 |
| Sensitivity | 0.713 | (0.637, 0.789) | 0.875 | (0.819, 0.931) | <0.0001 |
| Specificity | 0.712 | (0.677, 0.748) | 0.698 | (0.662, 0.734) | 0.3657 |
| PPV | 0.351 | (0.295, 0.408) | 0.388 | (0.333, 0.442) | 0.0330 |
| NPV | 0.919 | (0.895, 0.943) | 0.962 | (0.945, 0.980) | <0.0001 |
| AUPRC | 0.426 | (0.336, 0.517) | 0.533 | (0.427, 0.631) | 0.0020 |
| **Stress MBF** | | | | | |
| AUROC | 0.797 | (0.764, 0.830) | 0.835 | (0.805, 0.865) | 0.0023 |
| Sensitivity | 0.738 | (0.682, 0.795) | 0.742 | (0.686, 0.799) | 0.8759 |
| Specificity | 0.741 | (0.703, 0.778) | 0.790 | (0.756, 0.825) | 0.0108 |
| PPV | 0.558 | (0.503, 0.614) | 0.611 | (0.555, 0.668) | 0.0164 |
| NPV | 0.864 | (0.833, 0.896) | 0.874 | (0.844, 0.904) | 0.4566 |
| AUPRC | 0.637 | (0.566, 0.704) | 0.704 | (0.644, 0.755) | 0.0011 |
| **Stress TPD** | | | | | |
| AUROC | 0.783 | (0.747, 0.819) | 0.831 | (0.798, 0.865) | 0.0005 |
| Sensitivity | 0.680 | (0.616, 0.744) | 0.788 | (0.732, 0.844) | 0.0004 |
| Specificity | 0.759 | (0.723, 0.794) | 0.773 | (0.738, 0.808) | 0.3991 |
| PPV | 0.507 | (0.448, 0.567) | 0.559 | (0.502, 0.617) | 0.0127 |
| NPV | 0.866 | (0.836, 0.897) | 0.909 | (0.883, 0.935) | 0.0002 |

| | | | | | |
|---|---|---|---|---|---|
| AUPRC | 0.562 | (0.480, 0.633) | 0.673 | (0.605, 0.732) | <0.0001 |
| **LVEF** | | | | | |
| AUROC | 0.894 | (0.854, 0.934) | 0.949 | (0.927, 0.971) | 0.0005 |
| Sensitivity | 0.816 | (0.708, 0.925) | 0.918 | (0.842, 0.995) | 0.1317 |
| Specificity | 0.825 | (0.797, 0.853) | 0.883 | (0.859, 0.907) | <0.0001 |
| PPV | 0.244 | (0.178, 0.310) | 0.352 | (0.269, 0.434) | <0.0001 |
| NPV | 0.985 | (0.975, 0.995) | 0.994 | (0.987, 1.000) | 0.1058 |
| AUPRC | 0.435 | (0.292, 0.551) | 0.575 | (0.402, 0.736) | 0.0588 |
| **LVEF Reserve** | | | | | |
| AUROC | 0.813 | (0.746, 0.881) | 0.819 | (0.737, 0.902) | 0.8500 |
| Sensitivity | 0.833 | (0.684, 0.982) | 0.667 | (0.478, 0.855) | 0.1025 |
| Specificity | 0.661 | (0.627, 0.695) | 0.846 | (0.820, 0.872) | <0.0001 |
| PPV | 0.074 | (0.043, 0.106) | 0.124 | (0.067, 0.181) | 0.0078 |
| NPV | 0.992 | (0.984, 1.000) | 0.987 | (0.979, 0.996) | 0.2729 |
| AUPRC | 0.125 | (0.064, 0.256) | 0.222 | (0.088, 0.435) | 0.1889 |
| **Age (y)** | | | | | |
| MAE | 9.0 | (8.5, 9.5) | 7.6 | (7.2, 8.0) | <0.0001 |
| $R^2$ | 0.178 | (0.128, 0.229) | 0.382 | (0.327, 0.437) | <0.0001 |
| **BMI (kg/m$^2$)** | | | | | |
| MAE | 5.6 | (5.2, 5.9) | 4.4 | (4.2, 4.7) | <0.0001 |
| $R^2$ | 0.152 | (0.104, 0.200) | 0.454 | (0.401, 0.507) | <0.0001 |
| **Sex** | | | | | |
| AUROC | 0.812 | (0.782, 0.842) | 0.915 | (0.895, 0.935) | <0.0001 |
| Sensitivity | 0.758 | (0.717, 0.798) | 0.827 | (0.791, 0.862) | 0.0012 |
| Specificity | 0.742 | (0.694, 0.789) | 0.831 | (0.790, 0.872) | 0.0006 |
| PPV | 0.796 | (0.757, 0.835) | 0.867 | (0.834, 0.900) | <0.0001 |
| NPV | 0.697 | (0.648, 0.745) | 0.783 | (0.739, 0.826) | <0.0001 |
| AUPRC | 0.842 | (0.800, 0.880) | 0.933 | (0.911, 0.952) | <0.0001 |

*Supplementary Table S3: Holdout evaluation in the PTB-XL database. Task performance was compared with and without SSL pretraining. AUROC and Youden index are macro-averages over each multilabel task (Auto ECG Diagnosis, 44 labels; Auto ECG Rhythm, 12 labels; Auto ECG Form, 19 labels).*

| | No SSL Pretraining | | SSL Pretraining | | |
|---|---|---|---|---|---|
| | Estimate | 95% CI | Estimate | 95% CI | p-value |
| | | | **Auto ECG Diagnosis** | | |
| AUROC | 0.890 | (0.876, 0.904) | 0.925 | (0.916, 0.933) | <0.0001 |
| Youden Index | 0.531 | (0.492, 0.573) | 0.576 | (0.536, 0.615) | 0.1138 |
| | | | **Auto ECG Rhythm** | | |
| AUROC | 0.845 | (0.802, 0.887) | 0.974 | (0.958, 0.982) | <0.0001 |
| Youden Index | 0.440 | (0.341, 0.525) | 0.659 | (0.564, 0.750) | 0.0009 |
| | | | **Auto ECG Form** | | |
| AUROC | 0.806 | (0.782, 0.826) | 0.861 | (0.823, 0.890) | 0.0097 |
| Youden Index | 0.402 | (0.352, 0.449) | 0.502 | (0.434, 0.550) | 0.0089 |

*Supplementary Table S4: Analogous to Supplementary Table S2 but evaluated in the UM-PET holdout cohort (N=1031 patients).*

| | No SSL Pretraining | | SSL Pretraining | | |
|---|---|---|---|---|---|
| | Estimate | 95% CI | Estimate | 95% CI | p-value |
| **MFR** | | | | | |
| AUROC | 0.657 | (0.623, 0.690) | 0.763 | (0.735, 0.792) | <0.0001 |
| Sensitivity | 0.572 | (0.528, 0.615) | 0.743 | (0.705, 0.782) | <0.0001 |
| Specificity | 0.683 | (0.643, 0.722) | 0.657 | (0.617, 0.697) | 0.2593 |
| PPV | 0.625 | (0.580, 0.669) | 0.667 | (0.627, 0.706) | 0.0230 |
| NPV | 0.633 | (0.594, 0.673) | 0.735 | (0.695, 0.774) | <0.0001 |
| AUPRC | 0.643 | (0.587, 0.689) | 0.743 | (0.694, 0.784) | <0.0001 |
| **Rest MBF** | | | | | |
| AUROC | 0.767 | (0.735, 0.799) | 0.828 | (0.800, 0.855) | <0.0001 |
| Sensitivity | 0.690 | (0.639, 0.741) | 0.747 | (0.699, 0.795) | 0.0181 |
| Specificity | 0.738 | (0.706, 0.771) | 0.776 | (0.746, 0.807) | 0.0183 |
| PPV | 0.538 | (0.490, 0.587) | 0.596 | (0.548, 0.644) | 0.0015 |
| NPV | 0.843 | (0.815, 0.872) | 0.874 | (0.848, 0.900) | 0.0043 |
| AUPRC | 0.601 | (0.531, 0.666) | 0.691 | (0.628, 0.748) | <0.0001 |
| **Stress MBF** | | | | | |
| AUROC | 0.798 | (0.767, 0.830) | 0.866 | (0.840, 0.891) | <0.0001 |
| Sensitivity | 0.693 | (0.631, 0.755) | 0.767 | (0.711, 0.824) | 0.0025 |
| Specificity | 0.751 | (0.722, 0.781) | 0.803 | (0.775, 0.830) | 0.0001 |
| PPV | 0.423 | (0.372, 0.475) | 0.506 | (0.452, 0.560) | <0.0001 |
| NPV | 0.903 | (0.881, 0.925) | 0.929 | (0.910, 0.948) | 0.0003 |
| AUPRC | 0.507 | (0.433, 0.586) | 0.653 | (0.584, 0.715) | <0.0001 |
| **Stress TPD** | | | | | |
| AUROC | 0.709 | (0.676, 0.743) | 0.762 | (0.730, 0.794) | 0.0001 |
| Sensitivity | 0.667 | (0.617, 0.716) | 0.672 | (0.623, 0.721) | 0.8084 |
| Specificity | 0.666 | (0.631, 0.702) | 0.734 | (0.701, 0.767) | 0.0002 |
| PPV | 0.508 | (0.462, 0.553) | 0.566 | (0.518, 0.614) | 0.0008 |
| NPV | 0.795 | (0.762, 0.828) | 0.813 | (0.782, 0.844) | 0.1339 |
| AUPRC | 0.557 | (0.496, 0.618) | 0.666 | (0.607, 0.712) | <0.0001 |
| **LVEF** | | | | | |
| AUROC | 0.905 | (0.882, 0.929) | 0.955 | (0.942, 0.969) | <0.0001 |
| Sensitivity | 0.885 | (0.826, 0.944) | 0.920 | (0.870, 0.970) | 0.2850 |

| | | | | | |
|---|---|---|---|---|---|
| Specificity | 0.778 | (0.751, 0.805) | 0.844 | (0.821, 0.868) | <0.0001 |
| PPV | 0.329 | (0.276, 0.382) | 0.421 | (0.359, 0.483) | <0.0001 |
| NPV | 0.982 | (0.972, 0.992) | 0.989 | (0.981, 0.996) | 0.1960 |
| AUPRC | 0.569 | (0.453, 0.661) | 0.756 | (0.676, 0.819) | <0.0001 |
| **LVEF Reserve** | | | | | |
| AUROC | 0.779 | (0.673, 0.884) | 0.849 | (0.779, 0.919) | 0.1485 |
| Sensitivity | 0.708 | (0.526, 0.890) | 0.792 | (0.629, 0.954) | 0.3173 |
| Specificity | 0.726 | (0.698, 0.753) | 0.827 | (0.804, 0.851) | <0.0001 |
| PPV | 0.058 | (0.031, 0.085) | 0.098 | (0.056, 0.140) | 0.0007 |
| NPV | 0.991 | (0.984, 0.998) | 0.994 | (0.989, 0.999) | 0.1820 |
| AUPRC | 0.103 | (0.061, 0.176) | 0.100 | (0.062, 0.174) | 0.9361 |
| **Age (y)** | | | | | |
| MAE | 8.3 | (7.9, 8.7) | 6.8 | (6.5, 7.2) | <0.0001 |
| $R^2$ | 0.254 | (0.207, 0.300) | 0.487 | (0.442, 0.531) | <0.0001 |
| **BMI (kg/m$^2$)** | | | | | |
| MAE | 5.6 | (5.3, 5.9) | 4.5 | (4.3, 4.7) | <0.0001 |
| $R^2$ | 0.356 | (0.309, 0.404) | 0.578 | (0.538, 0.618) | <0.0001 |
| **Sex** | | | | | |
| AUROC | 0.895 | (0.876, 0.915) | 0.954 | (0.942, 0.966) | <0.0001 |
| Sensitivity | 0.849 | (0.820, 0.878) | 0.917 | (0.895, 0.939) | <0.0001 |
| Specificity | 0.773 | (0.734, 0.812) | 0.864 | (0.832, 0.896) | <0.0001 |
| PPV | 0.834 | (0.804, 0.863) | 0.900 | (0.876, 0.924) | <0.0001 |
| NPV | 0.793 | (0.755, 0.831) | 0.886 | (0.856, 0.916) | <0.0001 |
| AUPRC | 0.914 | (0.886, 0.935) | 0.962 | (0.946, 0.973) | <0.0001 |

*Supplementary Table S5: Benchmark comparison results for the LVEF prediction task (LVEF < 35%).*

| Classification | LVEF < 35% | | | | | |
|---|---|---|---|---|---|---|
| | Rest only ECG | | | | | |
| **Test data** | **UM-PET†** (present study) | **UM-phSPECT†** (present study) | **UM-exSPECT†** (present study) | **HM-PET†** (present study) | **UK Biobank†** (present study) | **UK Biobank*** (reference [101]) |
| N subjects | 1031 | 5102 | 1533 | 758 | 34,216 | 34,280 |
| Prevalence (%) | 11.0 | 3.5 | 0.5 | 6.9 | 0.3 | 0.3 |
| **Metrics** | | | | | | |
| AUROC | 0.950 | 0.943 | 0.935 | 0.949 | 0.833 | 0.720 |
| Sensitivity | 86.7 | 89.9 | 87.5 | 91.8 | 76.9 | 66.0 |
| Specificity | 88.9 | 86.9 | 97.4 | 88.3 | 83.5 | 66.0 |

† 1dViT + SSL pretraining (N=780,035) + fine-tuning (UM-PET training split, N=3126 ECGs)
* Deep CNN + supervised training (N=20,269 ECGs).

**[101] Hughes JW,** et al. Simple models vs. deep learning in detecting low ejection fraction from the electrocardiogram. *EHJ - Digital Health* 2024;5(4):427-434.

*Supplementary Table S6: Benchmark comparison results for the Age prediction task.*

| Regression | Age | | | | | | | |
|---|---|---|---|---|---|---|---|---|
| | Rest only ECG | | | | | | | |
| **Test data** | **UM-PET†** (present study) | **HM-PET†** (present study) | **UK Biobank†** (present study) | **UK Biobank *** (ref [102]) | **UK Biobank**** (ref [103]) | **PTB-XL†** (present study) | **PTB-XL ***** (ref [104]) | **PTB-XL ‡** (ref [105]) |
| N subjects | 1,031 | 758 | 66,670 | 36,349 | 54,190 | 2,164 | 2,164 | 21,837 |
| **Metrics** | | | | | | | | |
| MAE | 7.1 ± 5.4 y | 7.7 ± 5.6 y | 5.8 ± 4.3 y | 6.1 y | 7.7 y | 8.7 ± 6.3 y | 9.4 y | 8.8 / 8.3 y |
| $R^2$ | 0.452 | 0.382 | 0.225 | 0.281 | 0.209 | 0.652 | 0.590 | -- |

† 1dViT + SSL pretraining (N=780,035) + fine-tuning (UM-PET training split, N=3126 ECGs)
* Custom CNN + supervised training on (N=400,000 ECGs)
** Custom CNN + supervised training (N=230,000 ECGs)
*** `Resnet1D` + supervised training (N=1.5 million ECGs)
‡ Custom CNN / `Inception` models + supervised training (N=60,000 ECGs)

**[102] Libiseller-Egger J**, et al. Deep learning-derived cardiovascular age shares a genetic basis with other cardiac phenotypes. *Sci Rep* 2022;12(1):22625.
**[103] Evans S**, et al. Artificial intelligence age prediction using electrocardiogram data: Exploring biological age differences. *Heart Rhythm* 2024.
**[104] Hempel P,** et al. Towards explaining deep neural network-based heart age estimation. In: 2023 *IEEE EMBS Special Topic Conference on Data Science and Engineering in Healthcare, Medicine and Biology.;* 2023:41-42.
**[105] Singstad B-J,** Tavashi B. Using deep convolutional neural networks to predict patients age based on ECGs from an independent test cohort. *medRxiv* 2022:2022.10.03.22280640.

*Supplementary Table S7: Benchmark comparison results for the BMI prediction task (MAE=mean absolute error).*

| Regression | BMI | | | | |
|---|---|---|---|---|---|
| | Rest only ECG | | | | |
| **Test data** | **UM-PET†** (present study) | **HM-PET†** (present study) | **UK Biobank†** (present study) | **UK Biobank*** (ref [106]) | **PTB-XL†** (present study) |
| N subjects | 1031 | 758 | 65,025 | 42,386 | 678 |
| **Metrics** | | | | | |
| MAE | 4.7 ± 4.0 kg/m² | 4.4 ± 3.8 kg/m² | 4.6 ± 3.1 kg/m² | 2.9 ± 3.1 kg/m² | 3.4 ± 2.9 kg/m² |
| $R^2$ | 0.521 | 0.454 | 0.290 | 0.390 | 0.268 |

† 1dViT + SSL pretraining (N=780,035) + fine-tuning (UM-PET training split, N=3126 ECGs)
* `Resnet1D` + supervised training (N=500,000 ECGs)

**[106] Pastika L**, et al. Artificial intelligence-enhanced electrocardiography derived body mass index as a predictor of future cardiometabolic disease. *npj Digit. Med.* 2024;7(1):1-16.s

*Supplementary Table S8: Benchmark comparison results for the Sex prediction task.*

| **Classification** | **Sex** | | | | |
|---|---|---|---|---|---|
| | **Rest only ECG** | | | | |
| **Test data** | **UM-PET†** (present study) | **UK Biobank†** (present study) | **UK Biobank*** (ref [107]) | **PTB-XL†** (present study) | **PTB-XL**** (ref [108]) |
| N subjects | 1031 | 66,670 | 42,386 | 2198 | 2198 |
| **Metrics** | | | | | |
| AUROC | 0.945 | 0.921 | 0.971 | 0.848 | 0.920 |
| Sensitivity | 86.9 | 84.3 | -- | 73.6 | -- |
| Specificity | 86.4 | 84.4 | -- | 79.0 | -- |

† 1dViT + SSL pretraining (N=780,035) + fine-tuning (UM-PET training split, N=3126 ECGs)
* Custom CNN + supervised training (N=581,700 ECGs)
** Custom CNN + supervised training on 8 of 10 cross-validation folds (N=17,403) and tested on 10$^{th}$ holdout fold

**[107] Sau A,** et al. Artificial intelligence-enhanced electrocardiography for the identification of a sex-related cardiovascular risk continuum: a retrospective cohort study. *The Lancet Digital Health* 2025;7(3):e184–94.
**[108] Strodthoff N**, et al., Deep learning for ECG analysis: benchmarks and insights from PTB-XL. *IEEE Journal of Biomedical and Health Informatics* 2021;25(5):1519–28.

*Supplementary Table S9: Benchmark comparison results for three multi-label auto ECG interpretation tasks (Auto ECG Diagnosis, 44 labels; Auto ECG Rhythm, 12 labels; Auto ECG Form, 19 labels).*

| **Multilabel Classification** | **Auto ECG Interpretation** | | | | | |
|---|---|---|---|---|---|---|
| | **Rest only ECG** | | | | | |
| | **Diagnosis** | | **Rhythm** | | **Form** | |
| **Test data** | **PTB-XL†** (present study) | **PTB-XL*** (ref [108]) | **PTB-XL†** (present study) | **PTB-XL*** (ref [108]) | **PTB-XL†** (present study) | **PTB-XL*** (ref [108]) |
| N subjects | 2198 | 2198 | 2198 | 2198 | 2198 | 2198 |
| **Metrics** | | | | | | |
| Macro-averaged AUROC | 0.925 | 0.937 | 0.974 | 0.957 | 0.861 | 0.896 |

† 1dViT + SSL pretraining (N=780,035) + fine-tuning on 8 of 10 cross-validation folds (N=17,403 ECGs) and tested on 10th holdout fold
* Custom CNN + supervised training on 8 of 10 cross-validation folds (N=17,403 ECGs) and tested on 10th holdout fold

**[108] Strodthoff N**, et al., Deep learning for ECG analysis: benchmarks and insights from PTB-XL. *IEEE Journal of Biomedical and Health Informatics* 2021;25(5):1519–28.

*Supplementary Table S10: Confusion matrices for the predictions of classification tasks in the HM-PET database (N=758 patients).*

| | No SSL Pretraining | | | | SSL Pretraining | | | |
|---|---|---|---|---|---|---|---|---|
| | TP | TN | FP | FN | TP | TN | FP | FN |
| MFR | 248 | 259 | 127 | 123 | 278 | 256 | 130 | 94 |
| Rest MBF | 97 | 443 | 179 | 39 | 119 | 434 | 188 | 17 |
| Stress MBF | 172 | 389 | 136 | 61 | 173 | 415 | 110 | 60 |
| Stress TPD | 138 | 421 | 134 | 65 | 160 | 429 | 126 | 43 |
| LVEF | 40 | 585 | 124 | 9 | 45 | 626 | 83 | 4 |
| LVEF Reserve | 20 | 485 | 248 | 4 | 16 | 621 | 113 | 8 |
| Sex | 328 | 241 | 83 | 105 | 358 | 270 | 55 | 75 |

*Supplementary Table S11: Confusion matrices for the predictions of classification tasks in the UM-PET database (N=1031 patients).*

| | No SSL Pretraining | | | | SSL Pretraining | | | |
|---|---|---|---|---|---|---|---|---|
| | TP | TN | FP | FN | TP | TN | FP | FN |
| MFR | 283 | 366 | 170 | 212 | 368 | 352 | 184 | 127 |
| Rest MBF | 218 | 528 | 187 | 98 | 236 | 555 | 160 | 80 |
| Stress MBF | 149 | 613 | 203 | 66 | 165 | 655 | 161 | 50 |
| Stress TPD | 234 | 453 | 227 | 117 | 236 | 498 | 181 | 115 |
| LVEF | 100 | 714 | 204 | 13 | 104 | 775 | 143 | 9 |
| LVEF Reserve | 17 | 731 | 276 | 7 | 19 | 833 | 174 | 5 |
| Sex | 501 | 341 | 100 | 89 | 541 | 381 | 60 | 49 |

### Supplemental Figures

*Supplementary Figure S1: ECG-AI model architecture. (a) The one-dimensional transformer (1dViT) model; (b) Fusion model combining two 1dViT channels for rest and stress ECG input. (MLP, multilayer perceptron)*

*(a)*

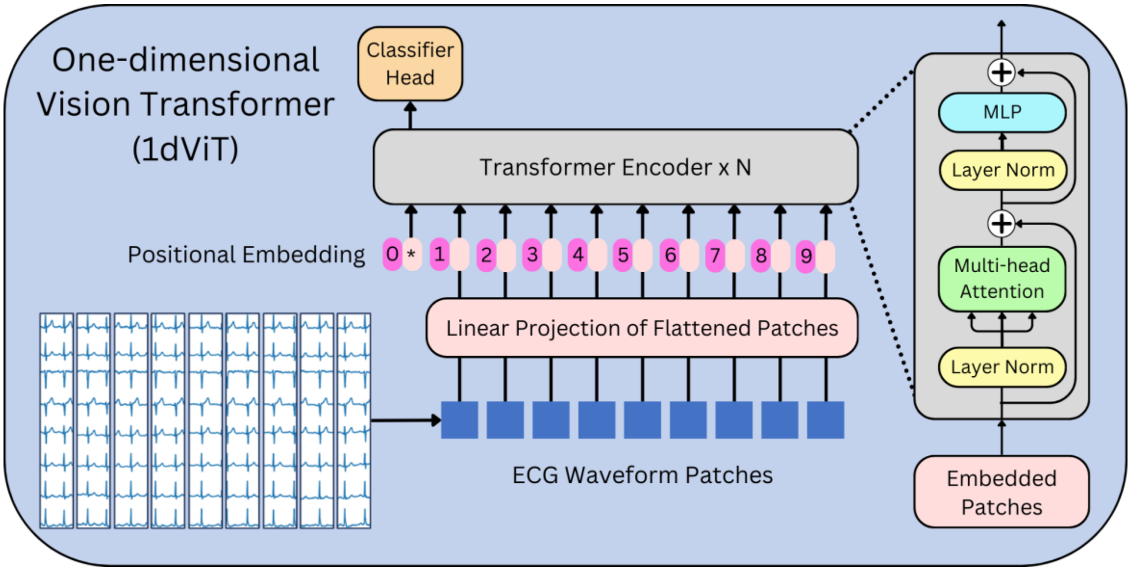


*(b)*

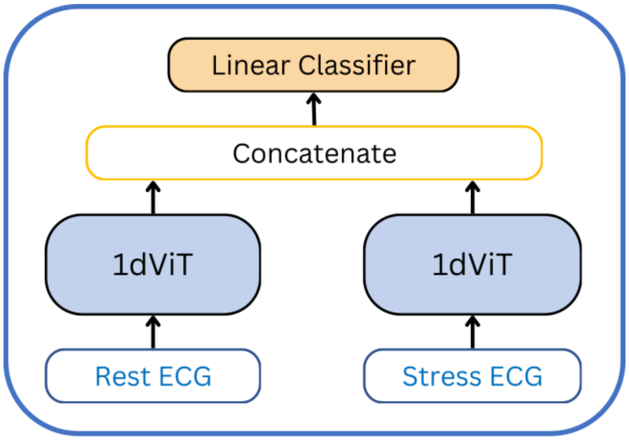

*Supplementary Figure S2: CONSORT diagram of UM-PET, UM-phSPECT, and UM-exSPECT MPI cohorts from the University of Michigan MPI stress testing registry.*

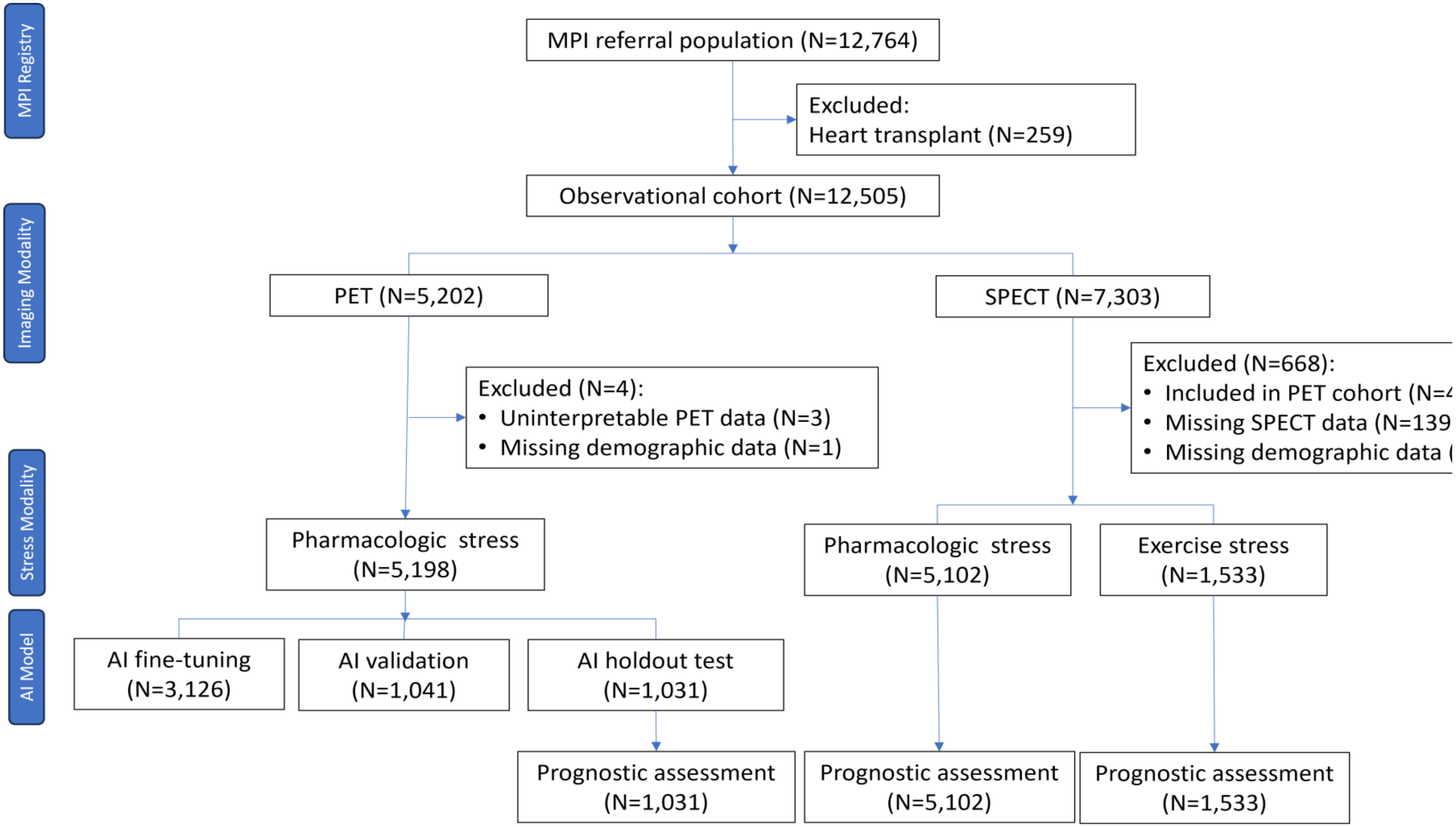

*Supplementary Figure S3: Analogous to Figure 2 in the main text but evaluated in the holdout UM-PET database. A complete summary is given in Supplementary Table S4.*

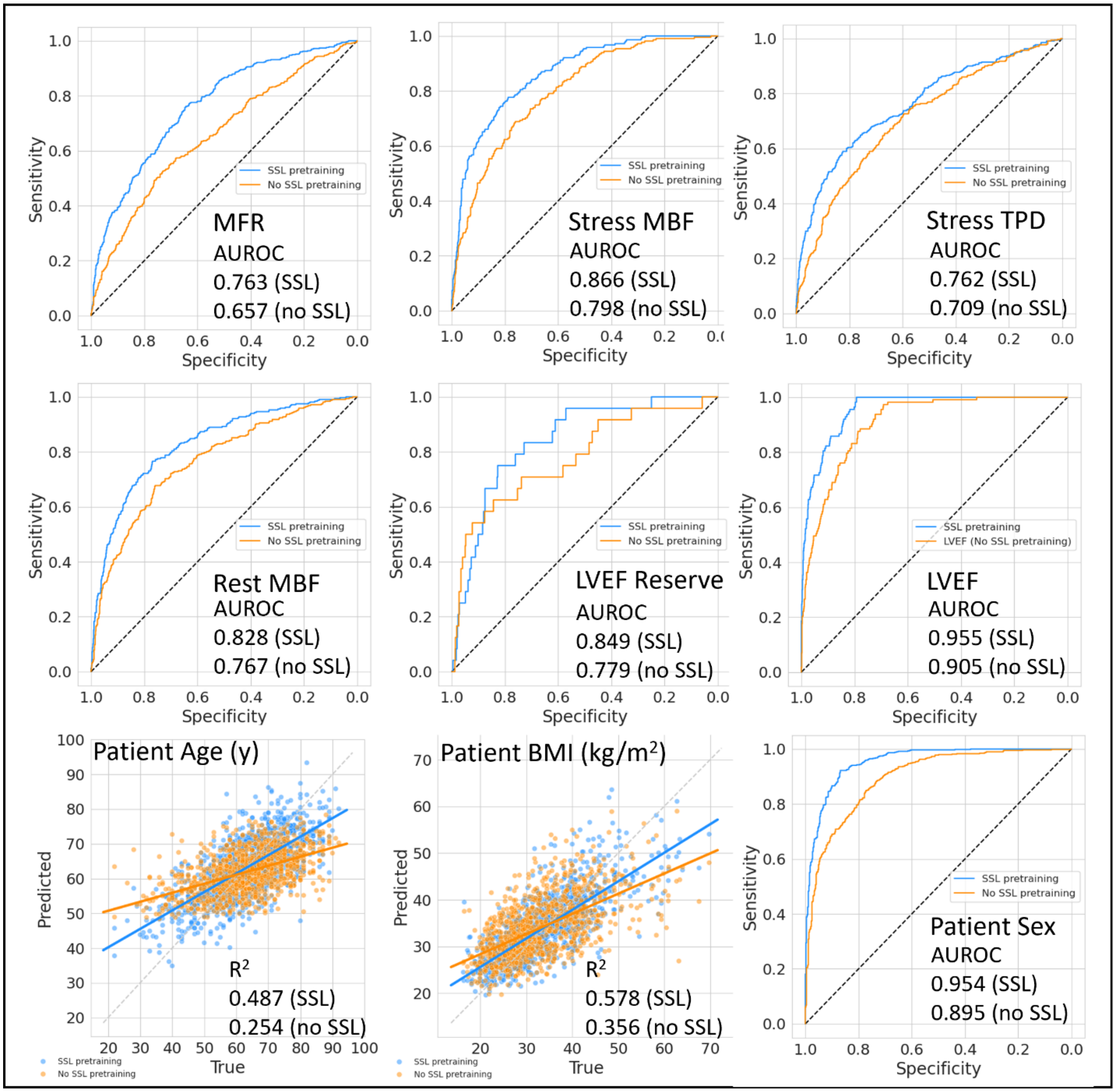

*Supplementary Figure S4: Comparison of model performance with and without SSL evaluated in the PTB-XL database. Model discrimination (AUROC) and calibration (ICI) are shown for each label of the multilabel predictions for (a) auto ECG diagnosis, (b) auto ECG rhythm, and (c) auto ECG form.*

*(a)*

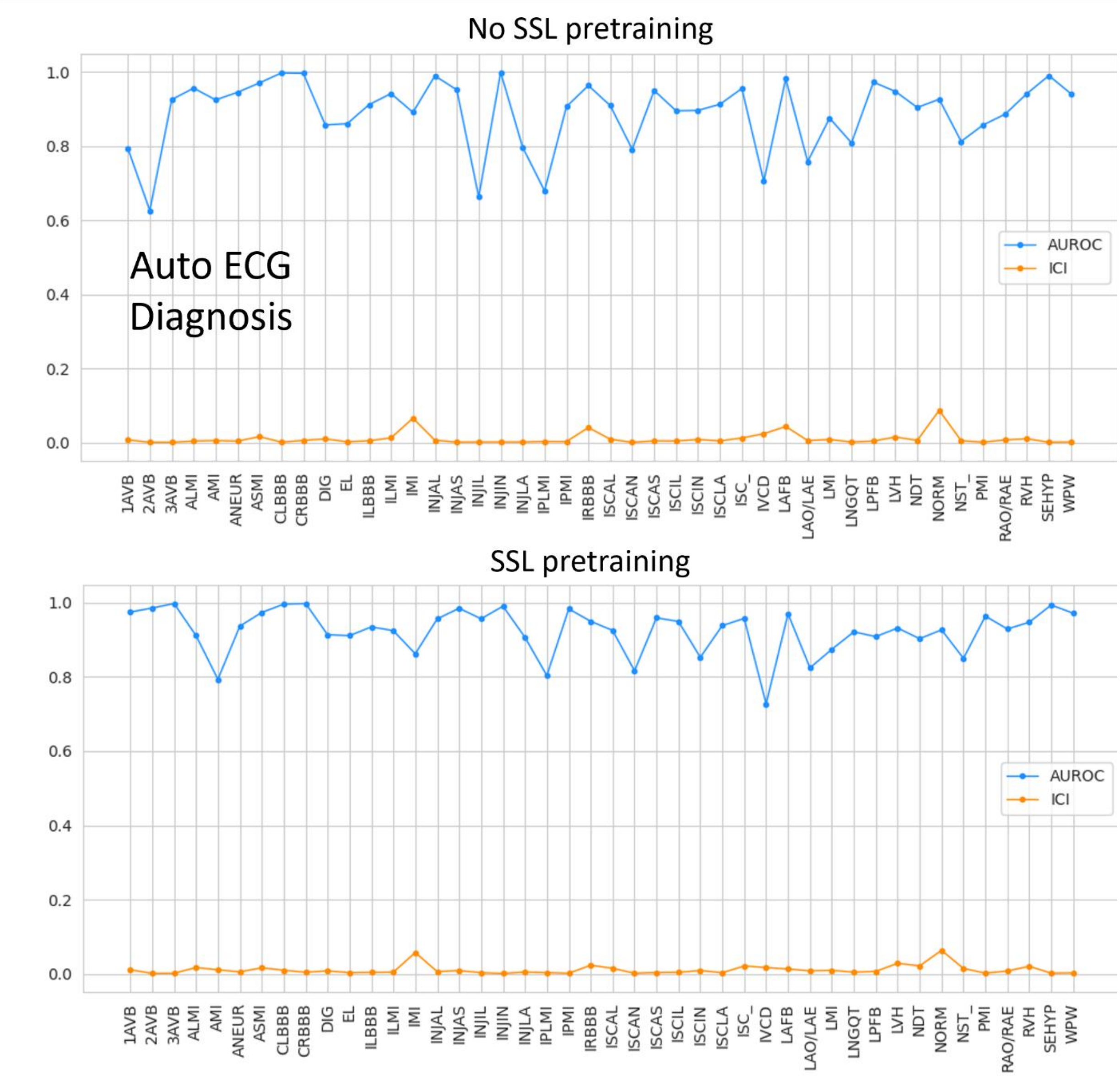


*(b)*

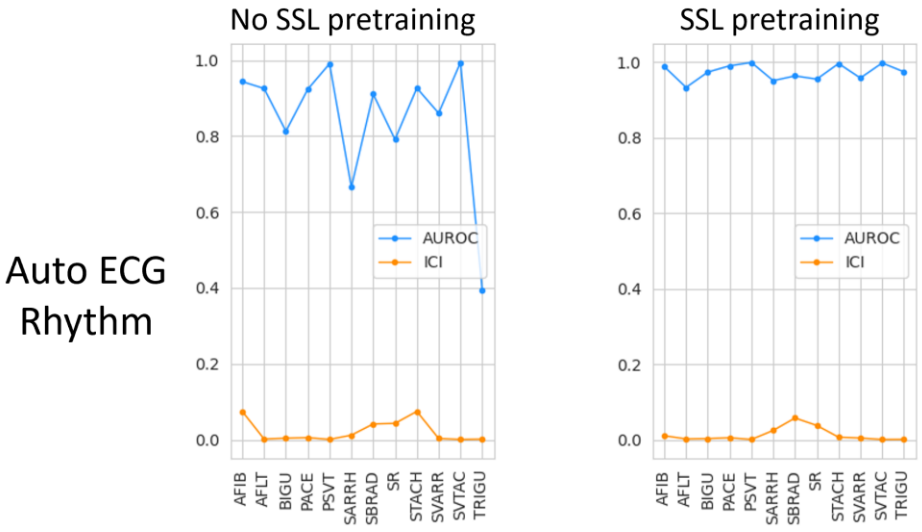
No SSL pretraining
SSL pretraining
Auto ECG Rhythm
AUROC
ICI
1.0
0.8
0.6
0.4
0.2
0.0
AFIB
AFLT
BIGU
PACE
PSVT
SARRH
SBRAD
SR
STACH
SVARR
SVTAC
TRIGU

*(c)*

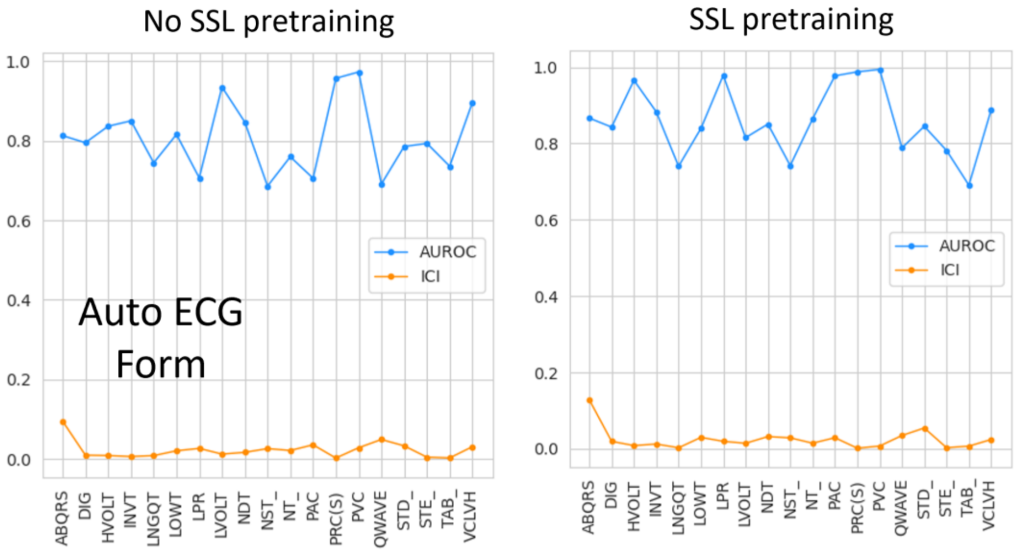
No SSL pretraining
SSL pretraining
Auto ECG Form
AUROC
ICI
1.0
0.8
0.6
0.4
0.2
0.0
ABQRS
DIG
HVOLT
INVT
LNGQT
LOWT
LPR
LVOLT
NDT
NST_
NT_
PAC
PRC(S)
PVC
QWAVE
STD_
STE_
TAB_
VCLVH

*Supplementary Figure S5: Examples of model calibration for two tasks, (a) MFR prediction, and (b) LVEF prediction. Label smoothing regularization was tuned during model fine-tuning by minimizing ICI and E90 which did not appreciably alter model discrimination (AUROC, Youden index) at the selected optimum.*

*(a)*

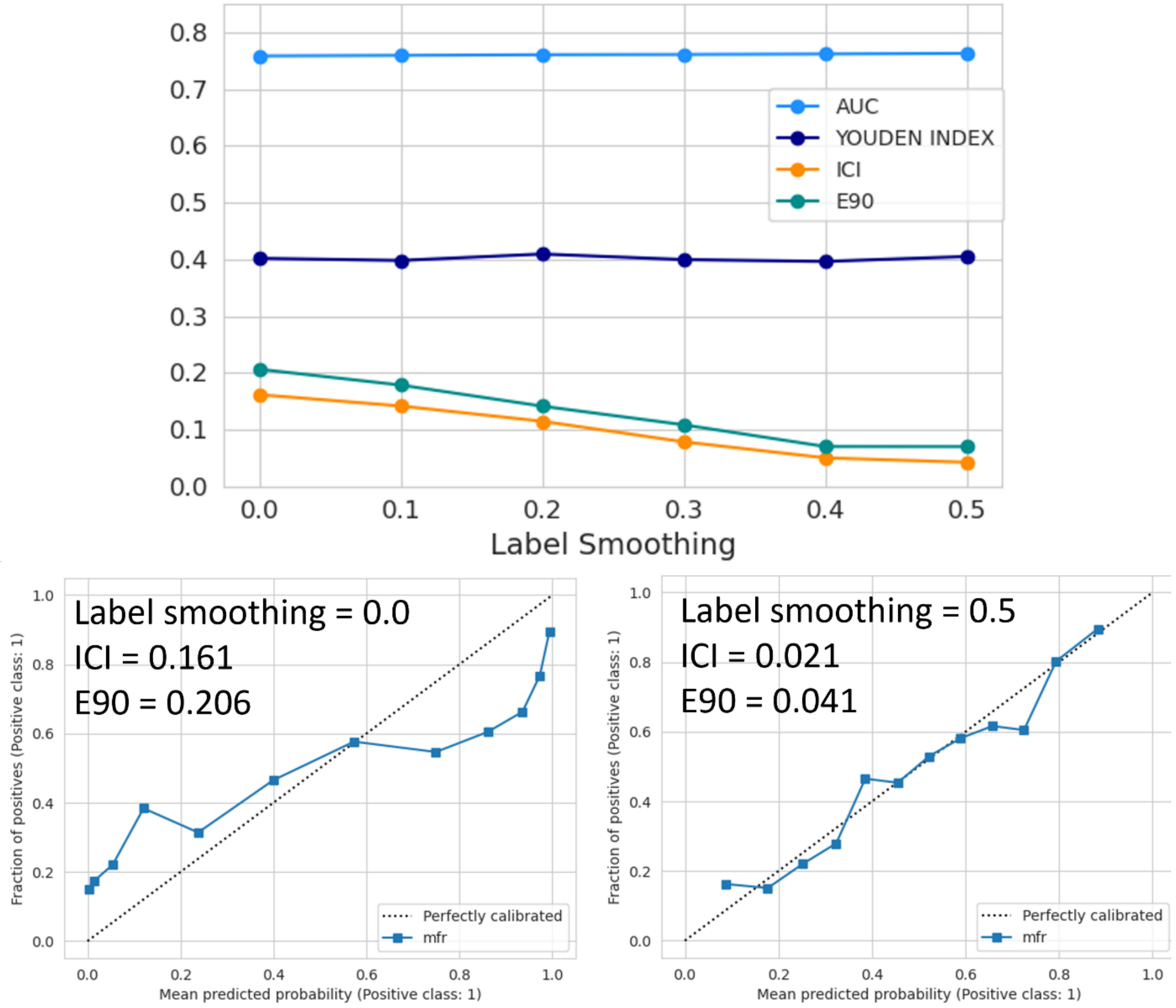


*(b)*

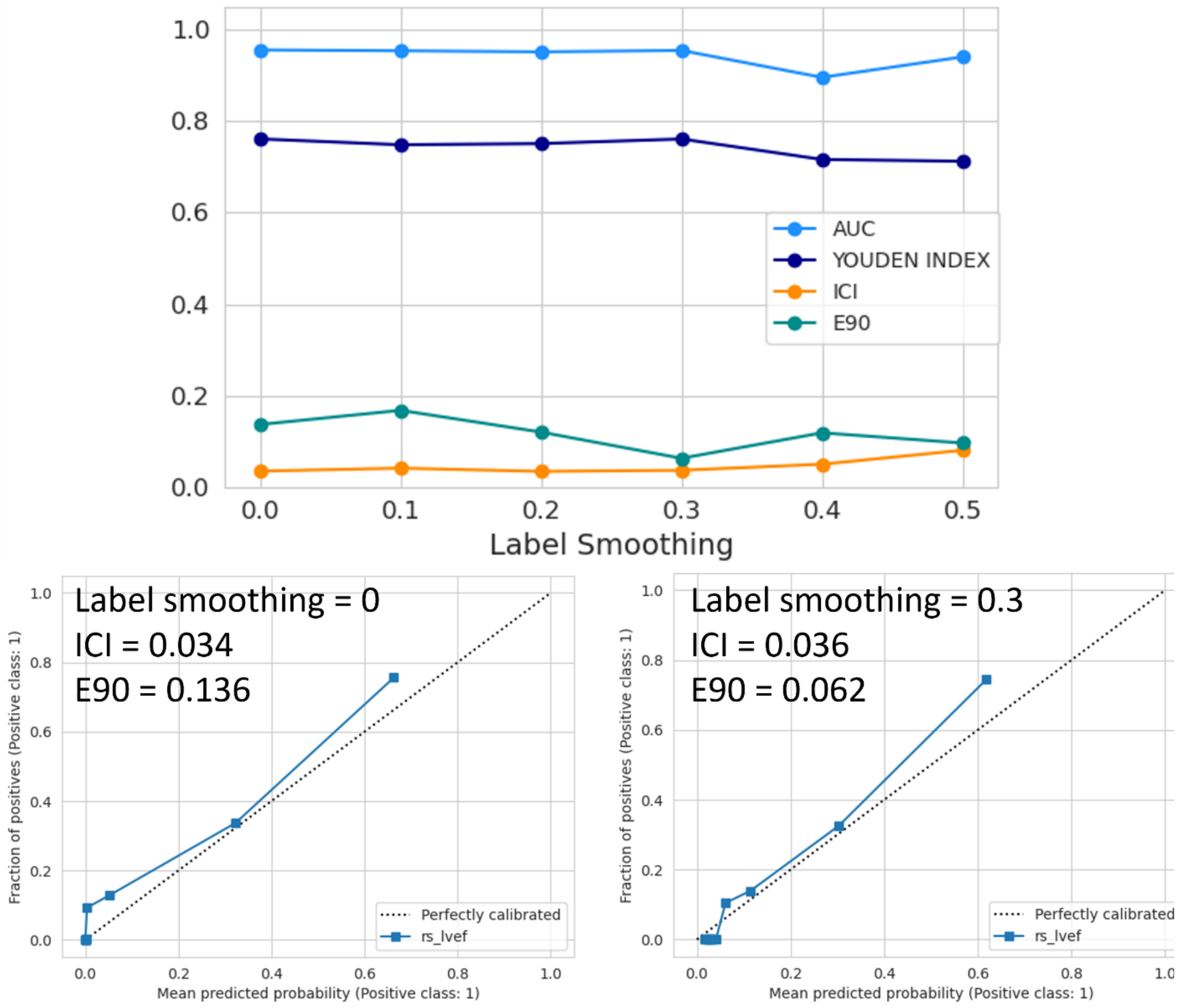
1.0
0.8
0.6
0.4
0.2
0.0
AUC
YOUDEN INDEX
ICI
E90
0.0 0.1 0.2 0.3 0.4 0.5
Label Smoothing
Label smoothing = 0
ICI = 0.034
E90 = 0.136
Fraction of positives (Positive class: 1)
Mean predicted probability (Positive class: 1)
Perfectly calibrated
rs_lvef
Label smoothing = 0.3
ICI = 0.036
E90 = 0.062
Fraction of positives (Positive class: 1)
Mean predicted probability (Positive class: 1)
Perfectly calibrated
rs_lvef

*Supplementary Figure S6: Subgroup analysis of SSL model performance in the UM-PET holdout database for (a) MFR prediction, (b) LVEF prediction, (c) stress TPD prediction, (d) rest MBF prediction, and (e) stress MBF prediction. Red dashed line indicates AUROC in the whole cohort.*

*(a)*

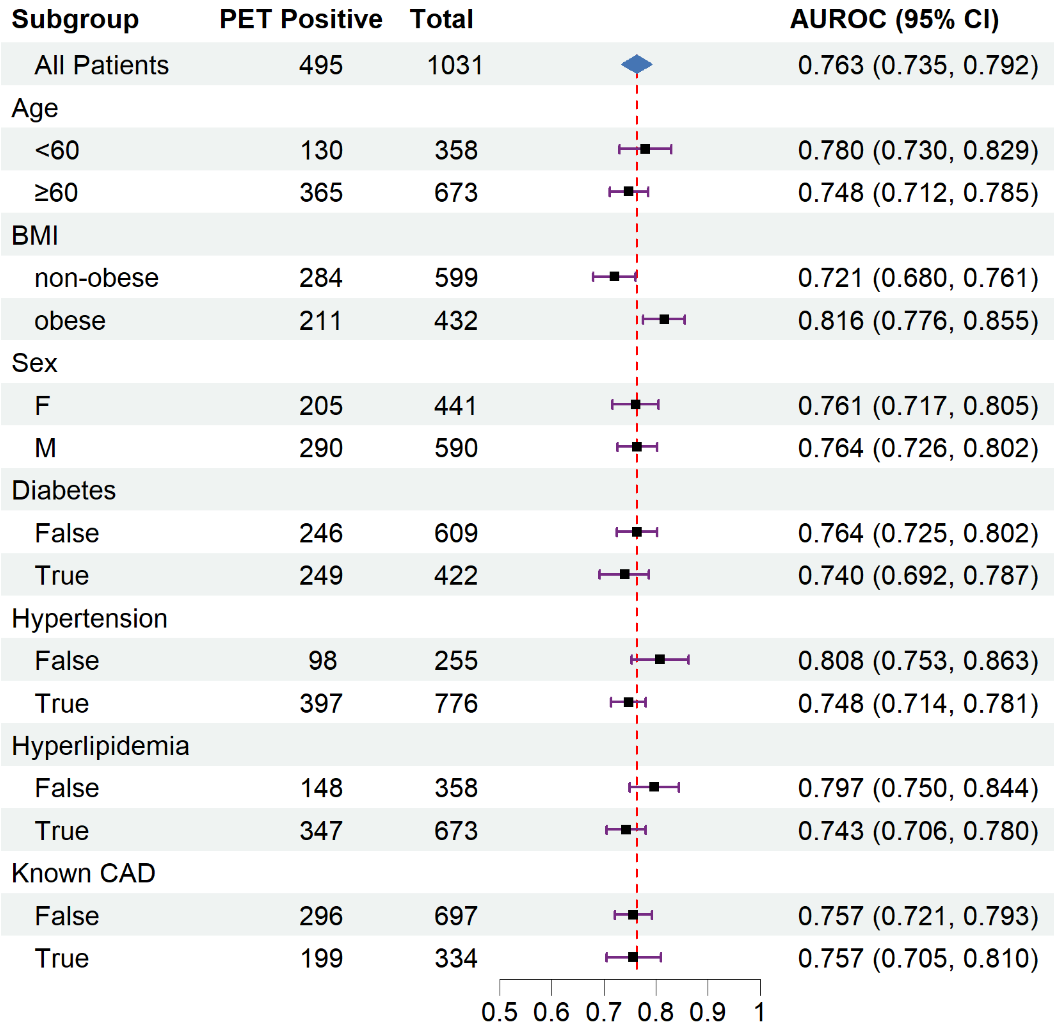


*(b)*

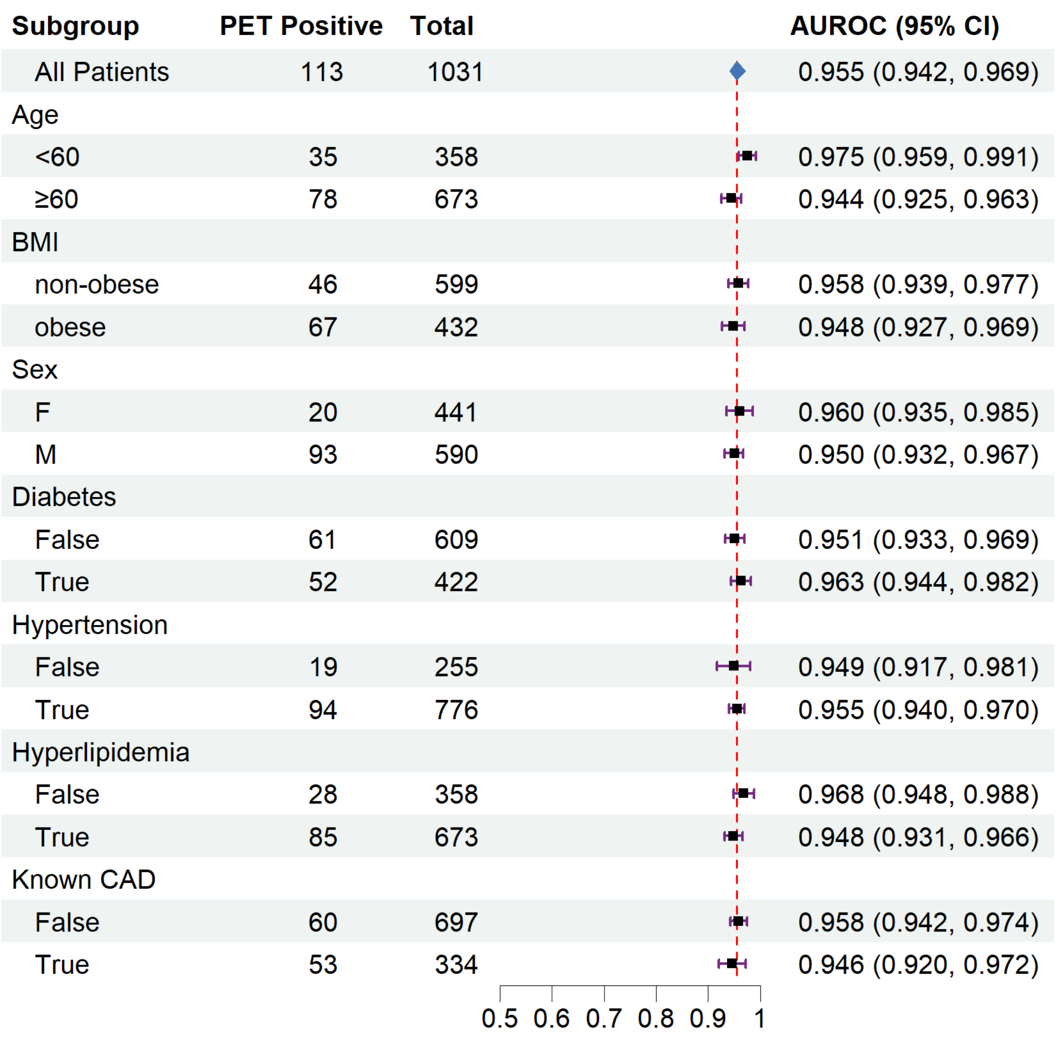
Subgroup PET Positive Total AUROC (95% CI)
All Patients 113 1031 0.955 (0.942, 0.969)
Age
<60 35 358 0.975 (0.959, 0.991)
≥60 78 673 0.944 (0.925, 0.963)
BMI
non-obese 46 599 0.958 (0.939, 0.977)
obese 67 432 0.948 (0.927, 0.969)
Sex
F 20 441 0.960 (0.935, 0.985)
M 93 590 0.950 (0.932, 0.967)
Diabetes
False 61 609 0.951 (0.933, 0.969)
True 52 422 0.963 (0.944, 0.982)
Hypertension
False 19 255 0.949 (0.917, 0.981)
True 94 776 0.955 (0.940, 0.970)
Hyperlipidemia
False 28 358 0.968 (0.948, 0.988)
True 85 673 0.948 (0.931, 0.966)
Known CAD
False 60 697 0.958 (0.942, 0.974)
True 53 334 0.946 (0.920, 0.972)
0.5 0.6 0.7 0.8 0.9 1

*(c)*

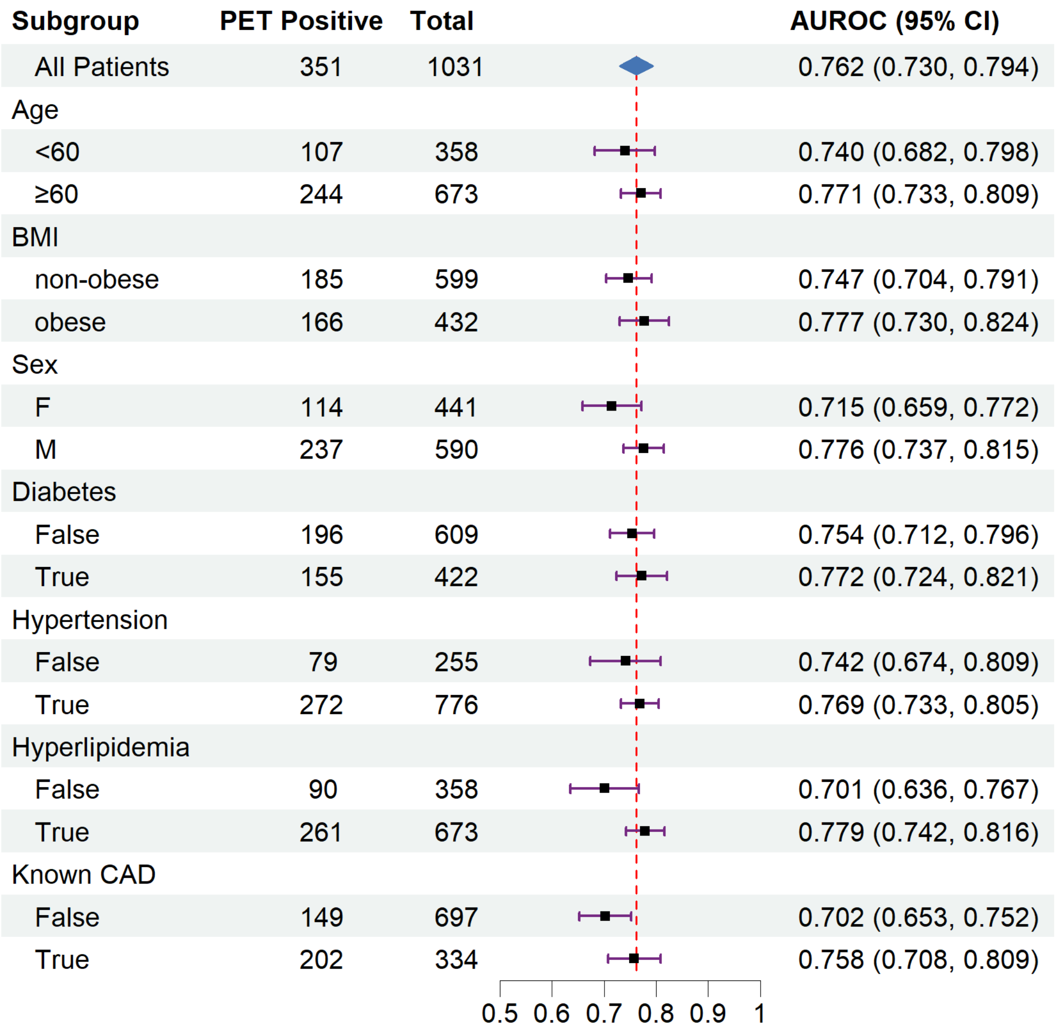
Subgroup
PET Positive
Total
AUROC (95% CI)
All Patients 351 1031 0.762 (0.730, 0.794)
Age
<60 107 358 0.740 (0.682, 0.798)
≥60 244 673 0.771 (0.733, 0.809)
BMI
non-obese 185 599 0.747 (0.704, 0.791)
obese 166 432 0.777 (0.730, 0.824)
Sex
F 114 441 0.715 (0.659, 0.772)
M 237 590 0.776 (0.737, 0.815)
Diabetes
False 196 609 0.754 (0.712, 0.796)
True 155 422 0.772 (0.724, 0.821)
Hypertension
False 79 255 0.742 (0.674, 0.809)
True 272 776 0.769 (0.733, 0.805)
Hyperlipidemia
False 90 358 0.701 (0.636, 0.767)
True 261 673 0.779 (0.742, 0.816)
Known CAD
False 149 697 0.702 (0.653, 0.752)
True 202 334 0.758 (0.708, 0.809)
0.5 0.6 0.7 0.8 0.9 1

*(d)*

| Subgroup | PET Positive | Total | AUROC (95% CI) |
|---|---|---|---|
| All Patients | 316 | 1031 | 0.828 (0.800, 0.855) |
| Age | | | |
| <60 | 121 | 358 | 0.796 (0.746, 0.846) |
| ≥60 | 195 | 673 | 0.847 (0.814, 0.879) |
| BMI | | | |
| non-obese | 210 | 599 | 0.808 (0.771, 0.844) |
| obese | 106 | 432 | 0.855 (0.810, 0.900) |
| Sex | | | |
| F | 201 | 441 | 0.796 (0.753, 0.838) |
| M | 115 | 590 | 0.807 (0.760, 0.854) |
| Diabetes | | | |
| False | 190 | 609 | 0.838 (0.804, 0.873) |
| True | 126 | 422 | 0.811 (0.765, 0.857) |
| Hypertension | | | |
| False | 84 | 255 | 0.825 (0.771, 0.879) |
| True | 232 | 776 | 0.828 (0.796, 0.861) |
| Hyperlipidemia | | | |
| False | 133 | 358 | 0.829 (0.784, 0.873) |
| True | 183 | 673 | 0.822 (0.786, 0.859) |
| Known CAD | | | |
| False | 249 | 697 | 0.824 (0.792, 0.857) |
| True | 67 | 334 | 0.810 (0.751, 0.869) |

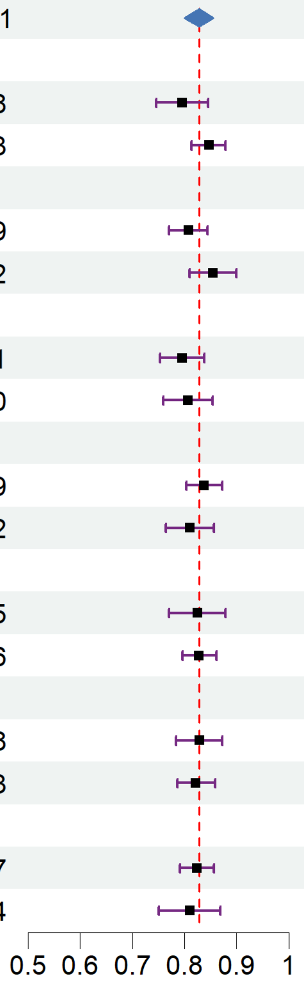


*(e)*

| Subgroup | PET Positive | Total | AUROC (95% CI) |
|---|---|---|---|
| All Patients | 215 | 1031 | 0.866 (0.840, 0.891) |
| Age | | | |
| <60 | 43 | 358 | 0.885 (0.837, 0.933) |
| ≥60 | 172 | 673 | 0.851 (0.819, 0.883) |
| BMI | | | |
| non-obese | 93 | 599 | 0.855 (0.814, 0.897) |
| obese | 122 | 432 | 0.867 (0.832, 0.902) |
| Sex | | | |
| F | 40 | 441 | 0.870 (0.816, 0.925) |
| M | 175 | 590 | 0.834 (0.799, 0.869) |
| Diabetes | | | |
| False | 93 | 609 | 0.853 (0.814, 0.892) |
| True | 122 | 422 | 0.872 (0.837, 0.908) |
| Hypertension | | | |
| False | 31 | 255 | 0.879 (0.825, 0.933) |
| True | 184 | 776 | 0.859 (0.829, 0.889) |
| Hyperlipidemia | | | |
| False | 35 | 358 | 0.849 (0.787, 0.912) |
| True | 180 | 673 | 0.863 (0.833, 0.893) |
| Known CAD | | | |
| False | 80 | 697 | 0.838 (0.796, 0.880) |
| True | 135 | 334 | 0.845 (0.804, 0.887) |

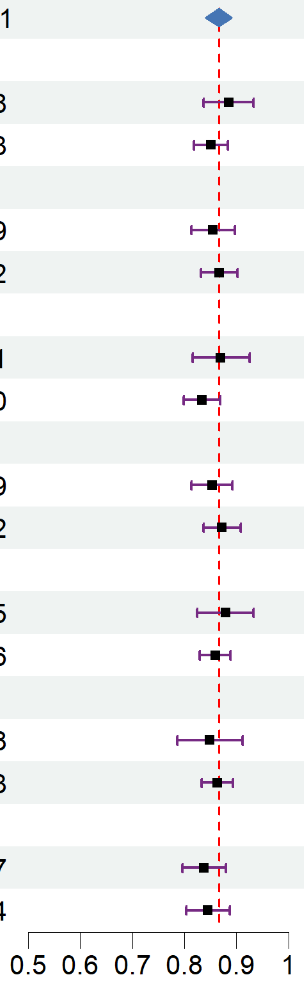

*Supplementary Figure S7: Subgroup analysis of SSL model performance in the HM-PET external database for (a) MFR prediction, (b) LVEF prediction, (c) stress TPD prediction, (d) rest MBF prediction, and (e) stress MBF prediction. Red dashed line indicates AUROC in the whole cohort.*

*(a)*

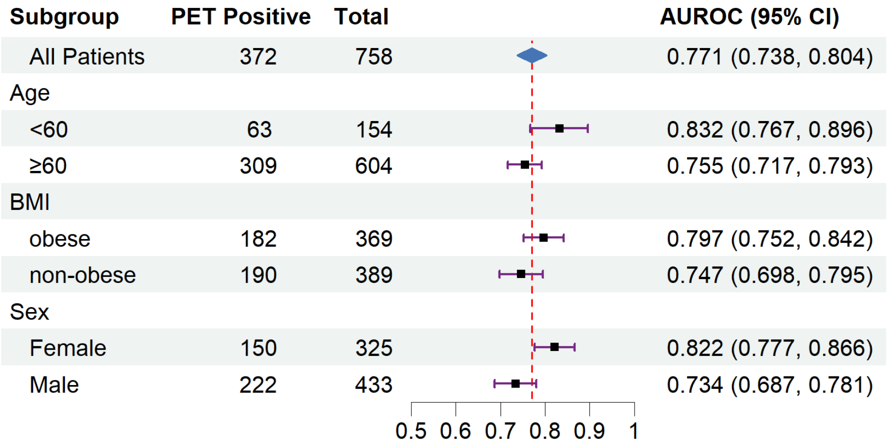


*(b)*

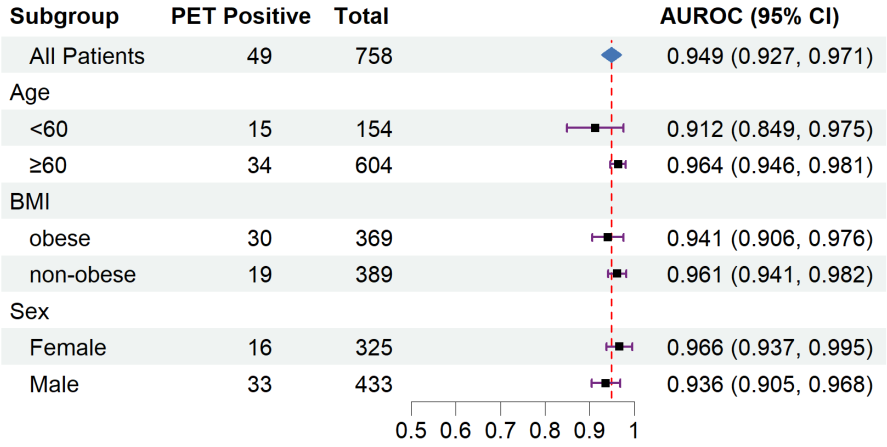


*(c)*

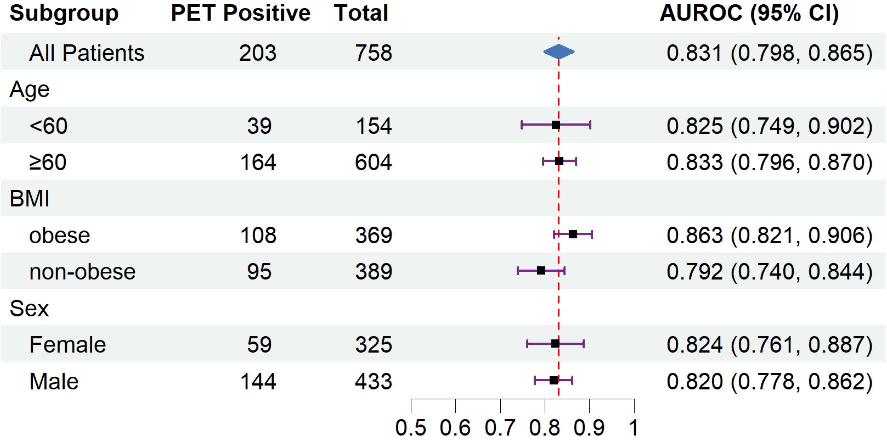

Subgroup | PET Positive | Total | AUROC (95% CI)
All Patients | 203 | 758 | 0.831 (0.798, 0.865)
Age
<60 | 39 | 154 | 0.825 (0.749, 0.902)
≥60 | 164 | 604 | 0.833 (0.796, 0.870)
BMI
obese | 108 | 369 | 0.863 (0.821, 0.906)
non-obese | 95 | 389 | 0.792 (0.740, 0.844)
Sex
Female | 59 | 325 | 0.824 (0.761, 0.887)
Male | 144 | 433 | 0.820 (0.778, 0.862)
0.5 0.6 0.7 0.8 0.9 1


*(d)*

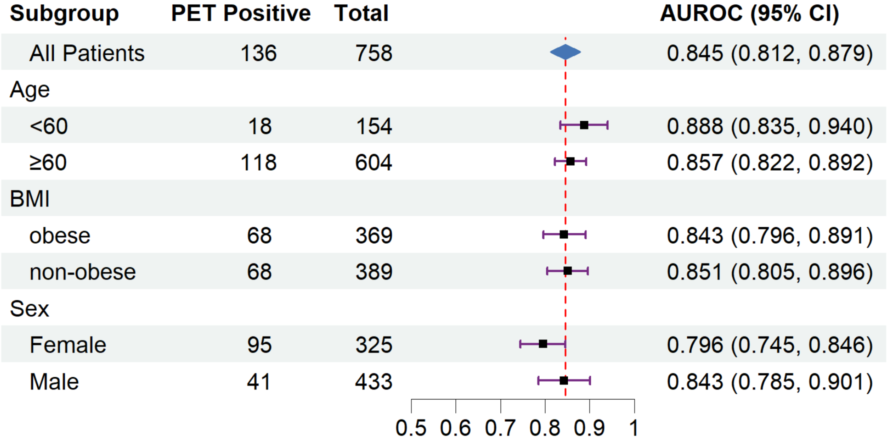

Subgroup | PET Positive | Total | AUROC (95% CI)
All Patients | 136 | 758 | 0.845 (0.812, 0.879)
Age
<60 | 18 | 154 | 0.888 (0.835, 0.940)
≥60 | 118 | 604 | 0.857 (0.822, 0.892)
BMI
obese | 68 | 369 | 0.843 (0.796, 0.891)
non-obese | 68 | 389 | 0.851 (0.805, 0.896)
Sex
Female | 95 | 325 | 0.796 (0.745, 0.846)
Male | 41 | 433 | 0.843 (0.785, 0.901)
0.5 0.6 0.7 0.8 0.9 1


*(e)*

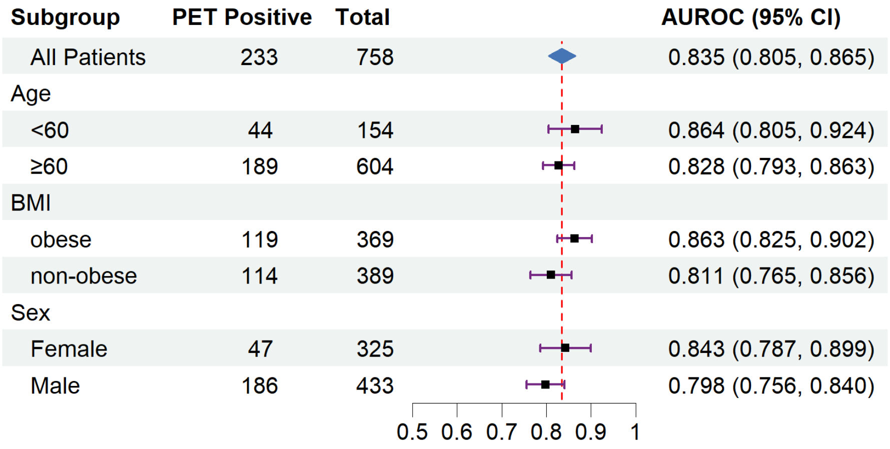
Subgroup
PET Positive
Total
AUROC (95% CI)
All Patients 233 758 0.835 (0.805, 0.865)
Age
<60 44 154 0.864 (0.805, 0.924)
≥60 189 604 0.828 (0.793, 0.863)
BMI
obese 119 369 0.863 (0.825, 0.902)
non-obese 114 389 0.811 (0.765, 0.856)
Sex
Female 47 325 0.843 (0.787, 0.899)
Male 186 433 0.798 (0.756, 0.840)
0.5 0.6 0.7 0.8 0.9 1

*Supplementary Figure S8: Subgroup analysis of SSL model performance in the UM-phSPECT out-of-distribution database for (a) LVEF prediction, (b) stress TPD prediction. Red dashed line indicates AUROC in the whole cohort.*

*(a)*

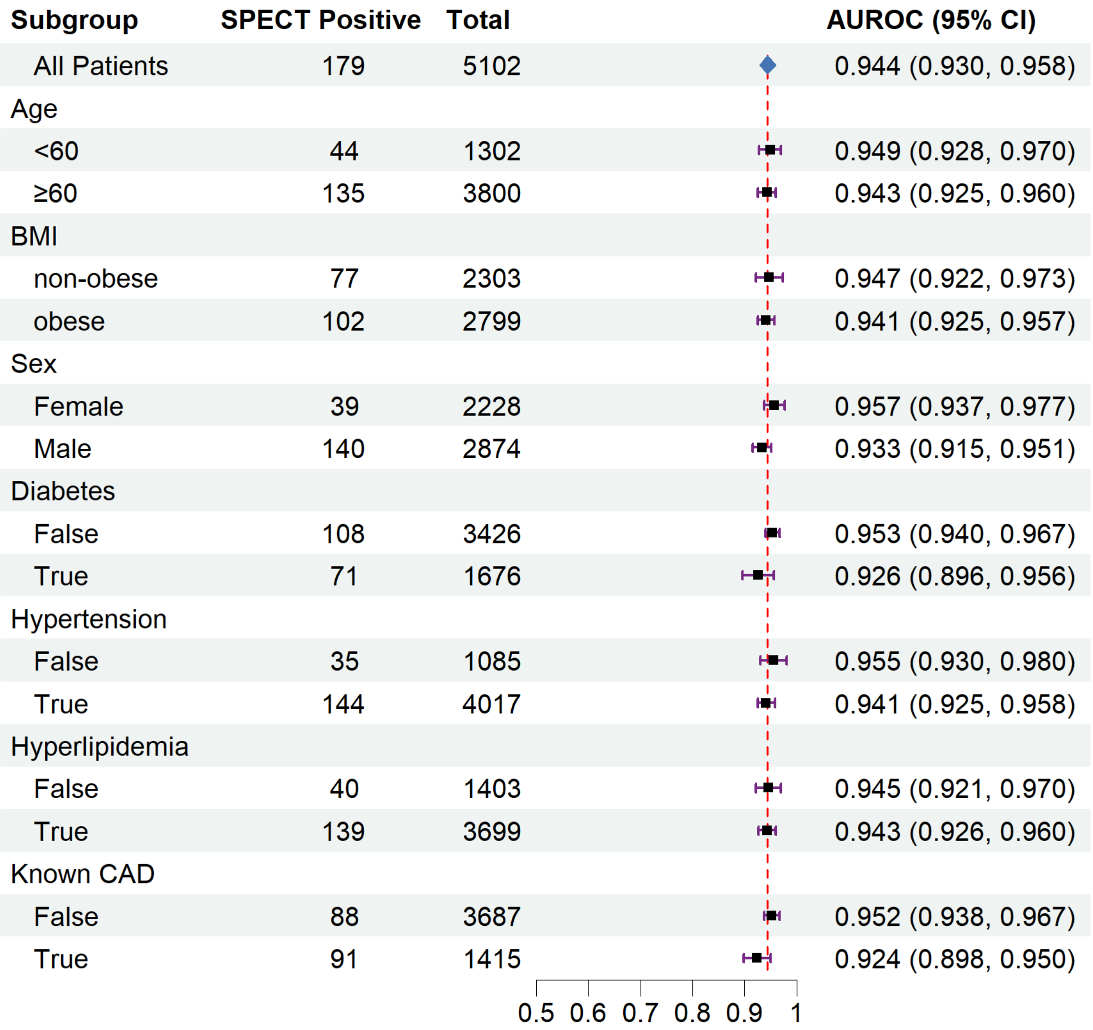


*(b)*

| Subgroup | SPECT Positive | Total | AUROC (95% CI) |
|---|---|---|---|
| All Patients | 1254 | 5102 | 0.744 (0.728, 0.760) |
| Age | | | |
| <60 | 271 | 1302 | 0.717 (0.683, 0.752) |
| ≥60 | 983 | 3800 | 0.751 (0.732, 0.769) |
| BMI | | | |
| non-obese | 552 | 2303 | 0.740 (0.715, 0.765) |
| obese | 702 | 2799 | 0.746 (0.724, 0.767) |
| Sex | | | |
| Female | 416 | 2228 | 0.709 (0.681, 0.738) |
| Male | 838 | 2874 | 0.753 (0.733, 0.773) |
| Diabetes | | | |
| False | 770 | 3426 | 0.734 (0.714, 0.755) |
| True | 484 | 1676 | 0.754 (0.728, 0.780) |
| Hypertension | | | |
| False | 215 | 1085 | 0.746 (0.707, 0.784) |
| True | 1039 | 4017 | 0.741 (0.723, 0.759) |
| Hyperlipidemia | | | |
| False | 246 | 1403 | 0.708 (0.672, 0.745) |
| True | 1008 | 3699 | 0.749 (0.731, 0.768) |
| Known CAD | | | |
| False | 609 | 3687 | 0.685 (0.661, 0.708) |
| True | 645 | 1415 | 0.764 (0.739, 0.788) |

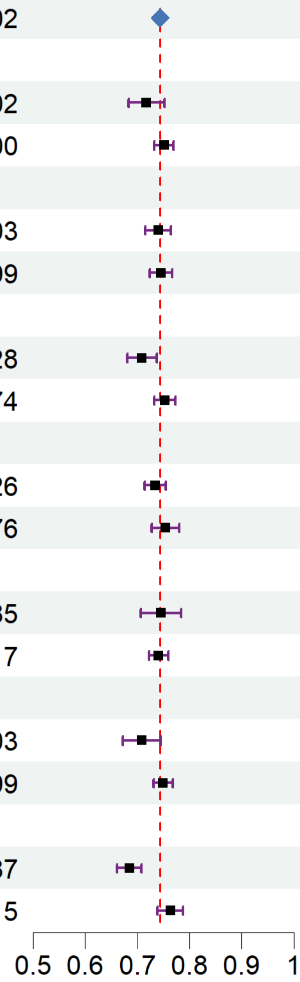

*Supplementary Figure S9: Subgroup analysis of SSL model performance in the UM-exSPECT out-of-distribution database for (a) LVEF prediction, (b) stress TPD prediction. Red dashed line indicates AUROC in the whole cohort.*

*(a)*

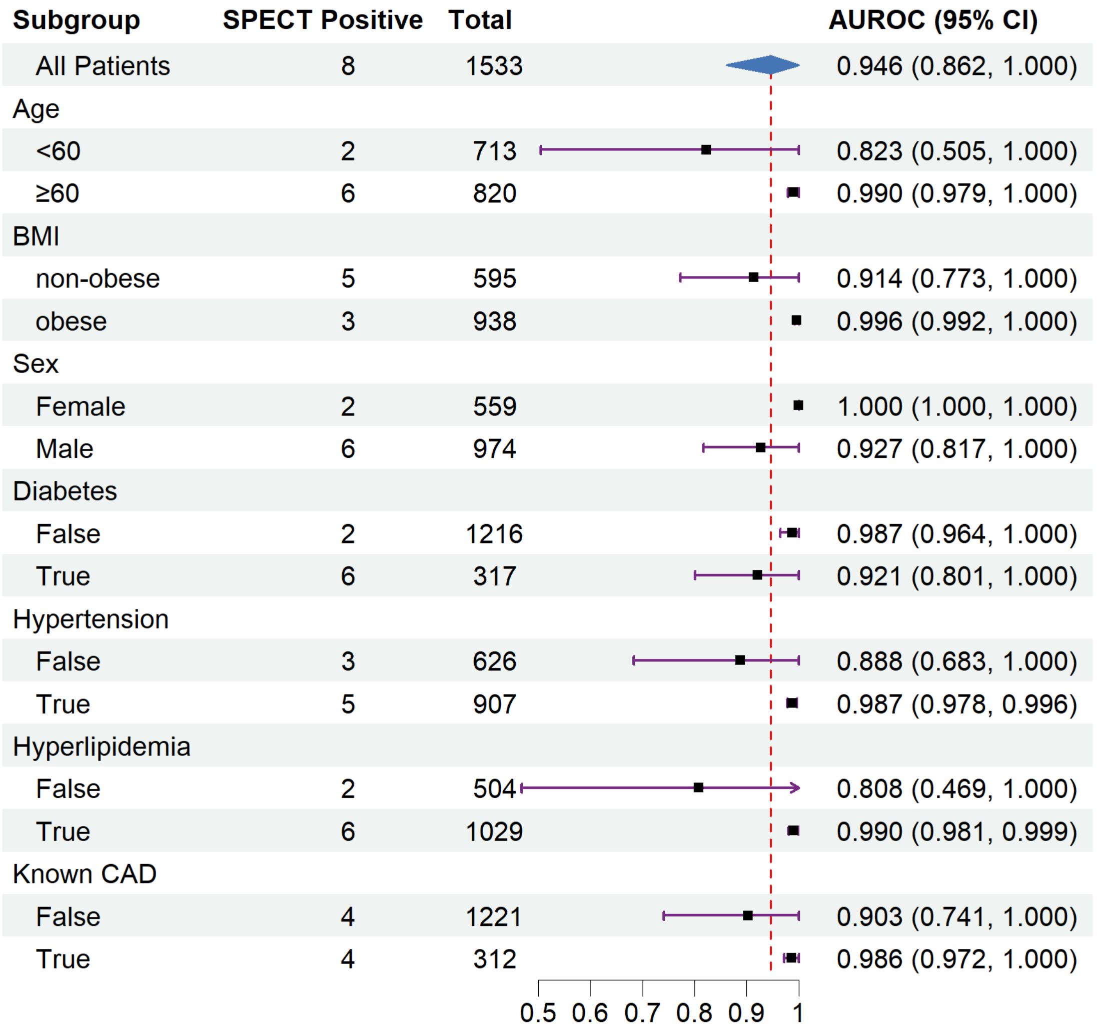


*(b)*

| Subgroup | SPECT Positive | Total | AUROC (95% CI) |
|---|---|---|---|
| All Patients | 203 | 1533 | 0.743 (0.704, 0.782) |
| Age | | | |
| <60 | 76 | 713 | 0.696 (0.626, 0.765) |
| ≥60 | 127 | 820 | 0.768 (0.721, 0.814) |
| BMI | | | |
| non-obese | 84 | 595 | 0.749 (0.686, 0.811) |
| obese | 119 | 938 | 0.740 (0.689, 0.791) |
| Sex | | | |
| Female | 51 | 559 | 0.710 (0.622, 0.797) |
| Male | 152 | 974 | 0.742 (0.697, 0.786) |
| Diabetes | | | |
| False | 145 | 1216 | 0.738 (0.692, 0.785) |
| True | 58 | 317 | 0.744 (0.667, 0.821) |
| Hypertension | | | |
| False | 49 | 626 | 0.636 (0.540, 0.733) |
| True | 154 | 907 | 0.765 (0.724, 0.807) |
| Hyperlipidemia | | | |
| False | 37 | 504 | 0.633 (0.519, 0.746) |
| True | 166 | 1029 | 0.763 (0.722, 0.804) |
| Known CAD | | | |
| False | 98 | 1221 | 0.704 (0.644, 0.765) |
| True | 105 | 312 | 0.744 (0.686, 0.801) |

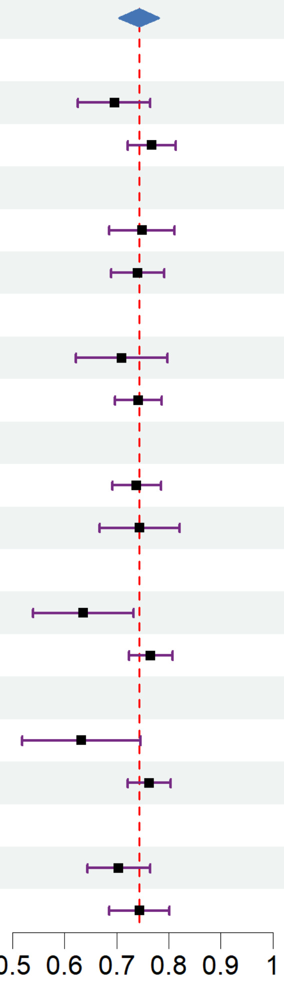

*Supplementary Figure S10: Subgroup analysis of SSL model performance in the UK Biobank external database for LVEF prediction. Red dashed line indicates AUROC in the whole cohort.*

| Subgroup | CMR Positive | Total | AUROC (95% CI) |
|---|---|---|---|
| All Patients | 91 | 34216 | 0.833 (0.776, 0.890) |
| Age | | | |
| <60 | 13 | 10662 | 0.706 (0.501, 0.910) |
| ≥60 | 78 | 23554 | 0.851 (0.795, 0.908) |
| BMI | | | |
| non-obese | 30 | 6456 | 0.841 (0.752, 0.930) |
| obese | 61 | 27760 | 0.824 (0.750, 0.898) |
| Sex | | | |
| F | 21 | 17806 | 0.706 (0.570, 0.843) |
| M | 70 | 16410 | 0.867 (0.806, 0.927) |
| Diabetes | | | |
| False | 89 | 33969 | 0.830 (0.772, 0.888) |
| True | 2 | 247 | 0.976 (0.946, 1.000) |
| Hypertension | | | |
| False | 54 | 26367 | 0.779 (0.697, 0.862) |
| True | 37 | 7849 | 0.909 (0.848, 0.971) |
| Hyperlipidemia | | | |
| False | 56 | 26369 | 0.796 (0.716, 0.875) |
| True | 35 | 7847 | 0.888 (0.816, 0.961) |
| Known CAD | | | |
| False | 78 | 33482 | 0.812 (0.747, 0.877) |
| True | 13 | 734 | 0.929 (0.886, 0.972) |

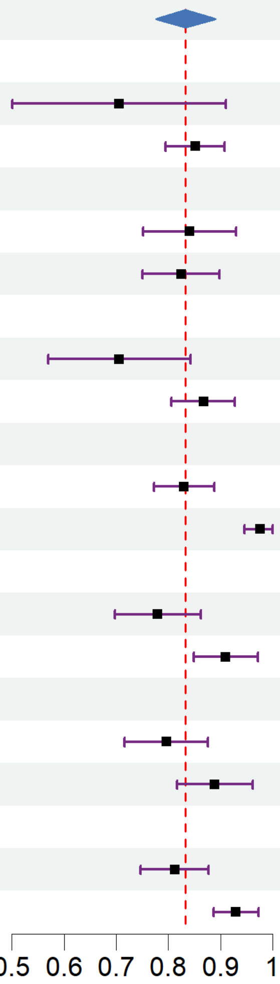

*Supplementary Figure S11: Learning rate sweeps evaluated in the UM-PET holdout test cohort for three prediction tasks: (a) MFR, (b) LVEF, (c) rest MBF.*

*(a)*

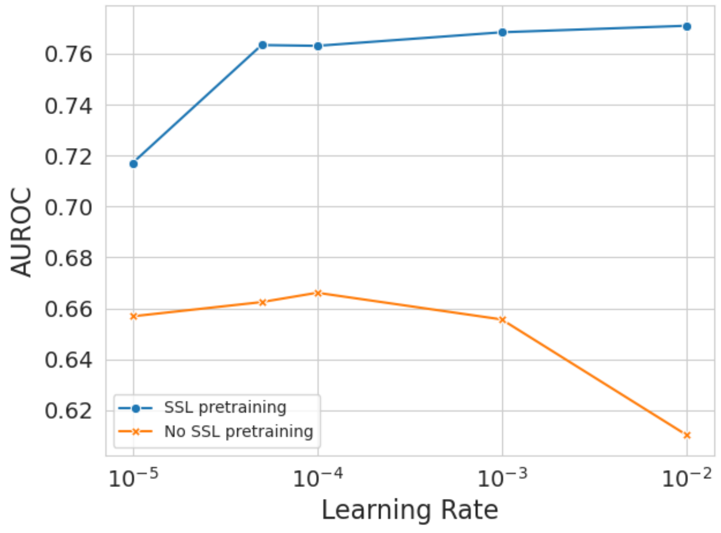


*(b)*

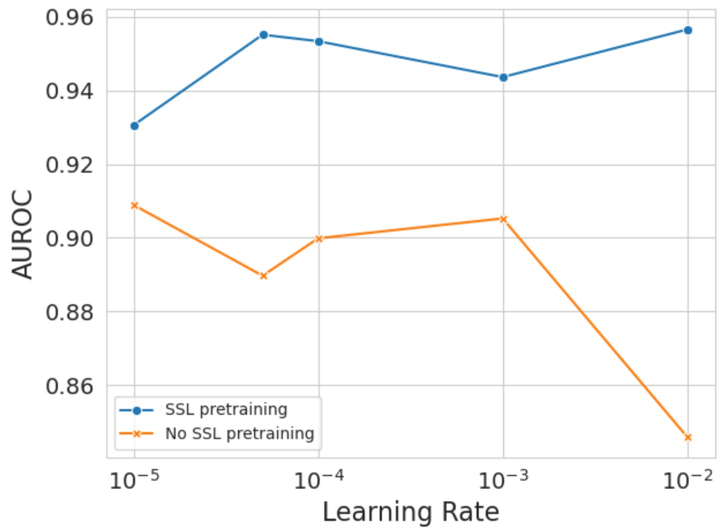

(c)

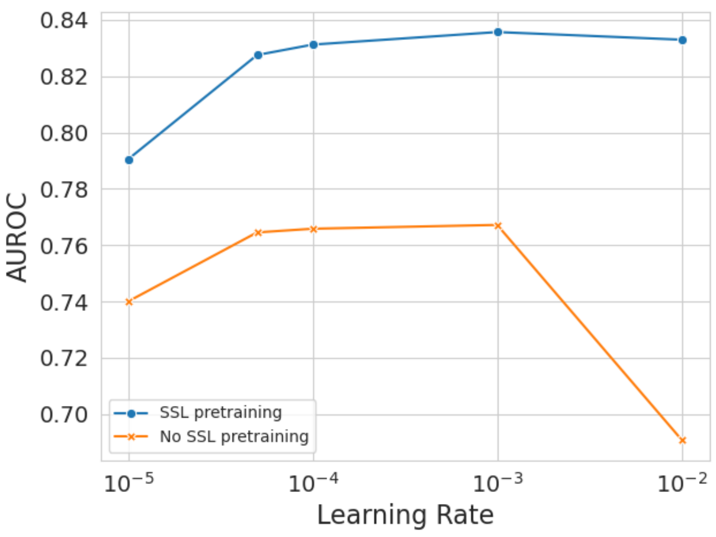

0.84
0.82
0.80
0.78
0.76
0.74
0.72
0.70
AUROC
SSL pretraining
No SSL pretraining
$10^{-5}$
$10^{-4}$
$10^{-3}$
$10^{-2}$
Learning Rate

*Supplementary Figure S12: Event rates across three cohorts from the University of Michigan. Differences in referral patterns, patient characteristics and risk factor profiles between cohorts (Table 3 and Supplementary Table S1) resulted in significant differences in clinical outcomes.*

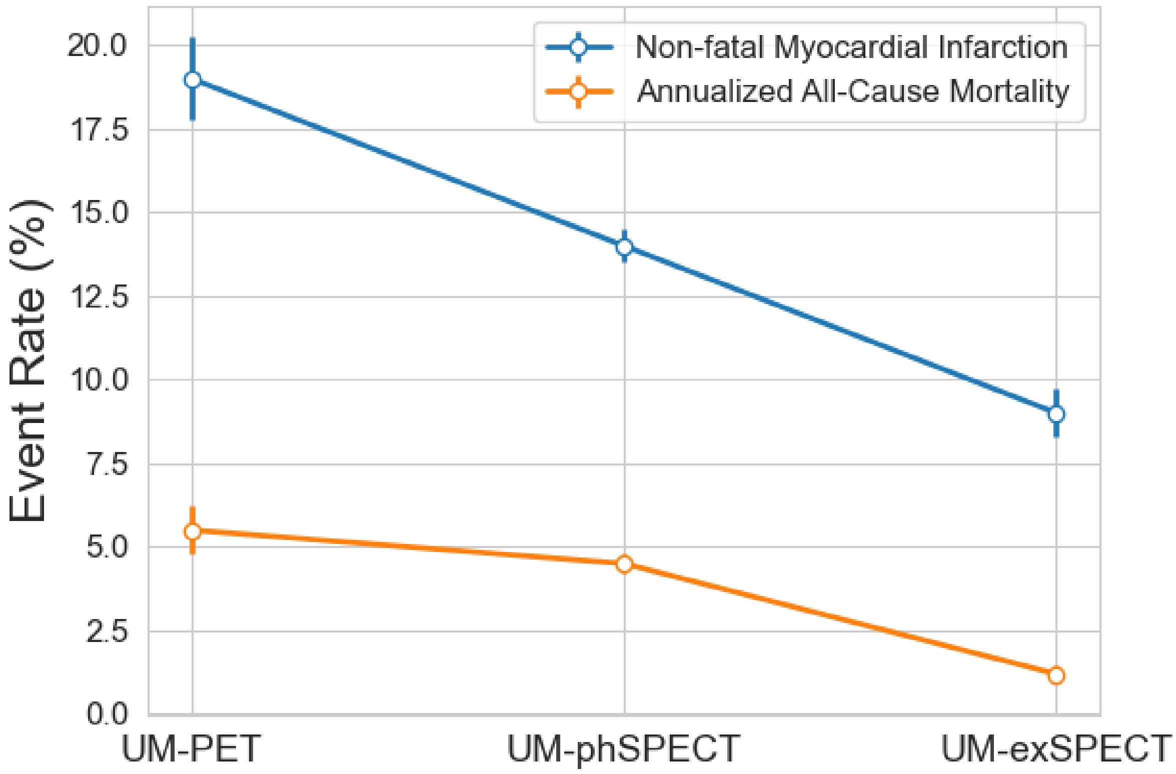

*Supplementary Figure S13: Additional examples of model calibration for (a) rest MBF and (b) stress TPD prediction tasks.*

*(a)*

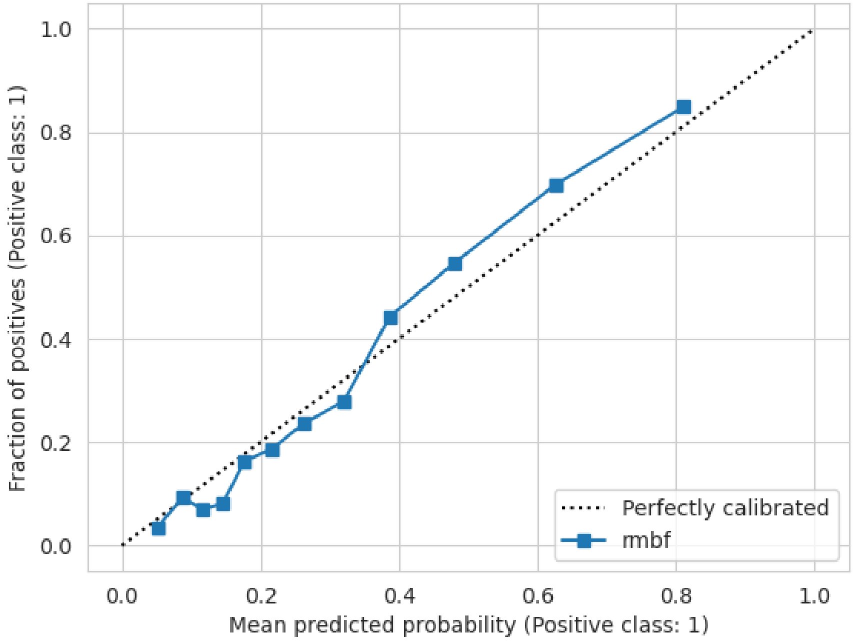


*(b)*

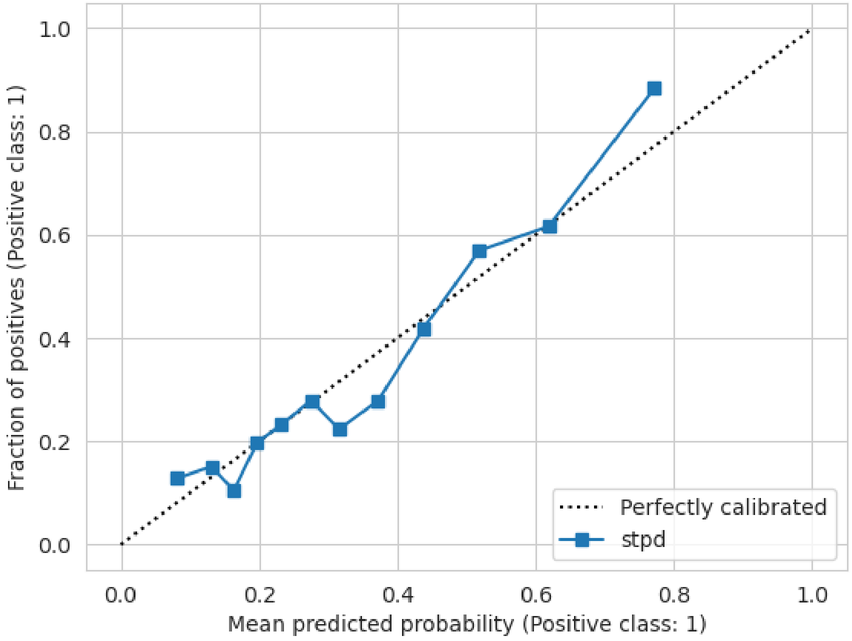

*Supplementary Figure S14: 2D projections of the SSL-pretrained 1dViT feature map without fine-tuning. The feature map was extracted from the last transformer block before the final classifier head computed using the UM-PET training split and displayed with (a) MFR labels and (b) LVEF labels. The SSL pretraining generated five clusters with a distinct global structure but without clear separation of labels in (a) but some separation in (b).*

*(a)*

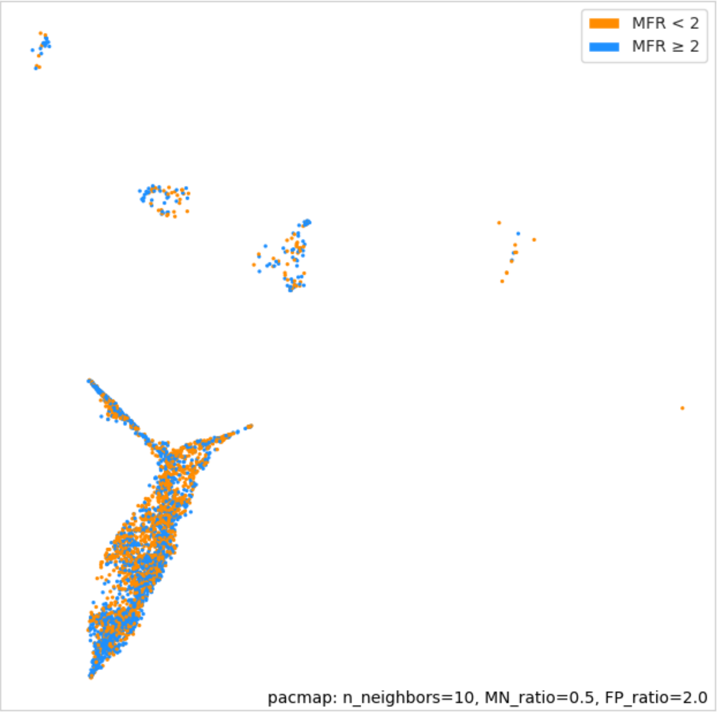


*(b)*

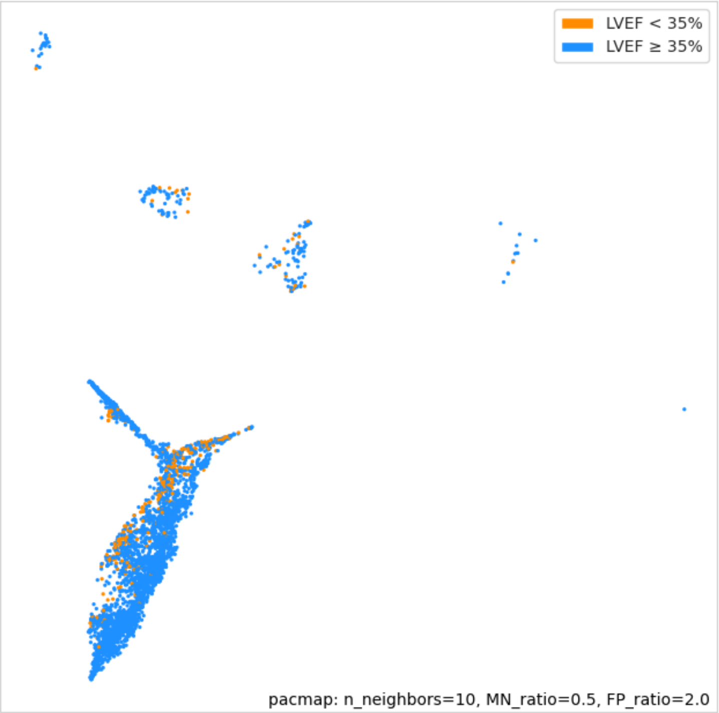

*Supplementary Figure S15: 2D projections of feature maps after fine-tuning for the MFR task on the UM-PET training split. (a) Feature map of stress-rest ECG input to the SSL-pretrained dual 1dViT; (b) feature map of rest-only ECG input to the SSL-pretrained 1dViT; (c) feature map of stress-rest ECG input to the de novo dual 1dViT (no SSL). Fine-tuning the SSL model generally had the effect of collapsing 3-4 of the smaller clusters and shifting labels within the larger global structure to facilitate somewhat better separation. De novo supervised training (without SSL) generated feature maps with more diffuse, less clearly defined global structure and notably poorer label separation.*

*(a)*

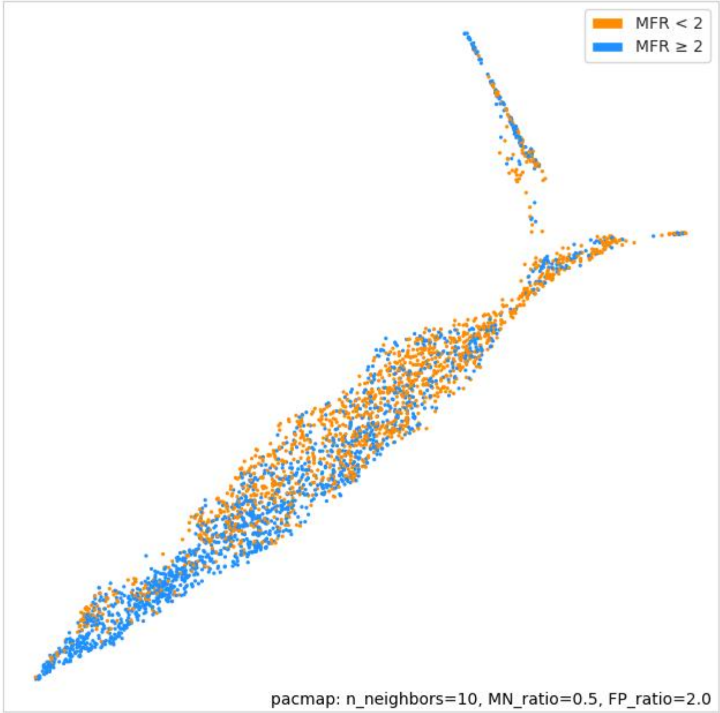

*(b)*

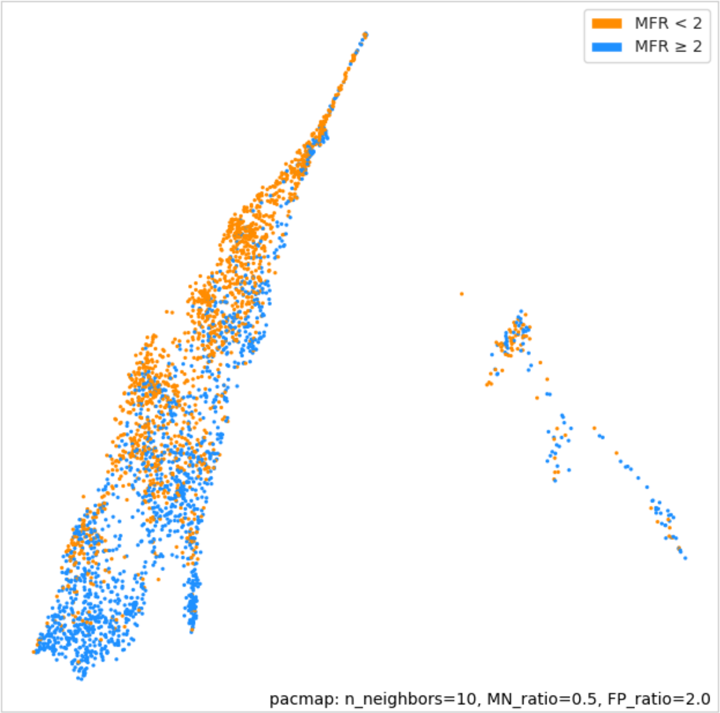
MFR < 2
MFR ≥ 2
pacmap: n_neighbors=10, MN_ratio=0.5, FP_ratio=2.0

*(c)*

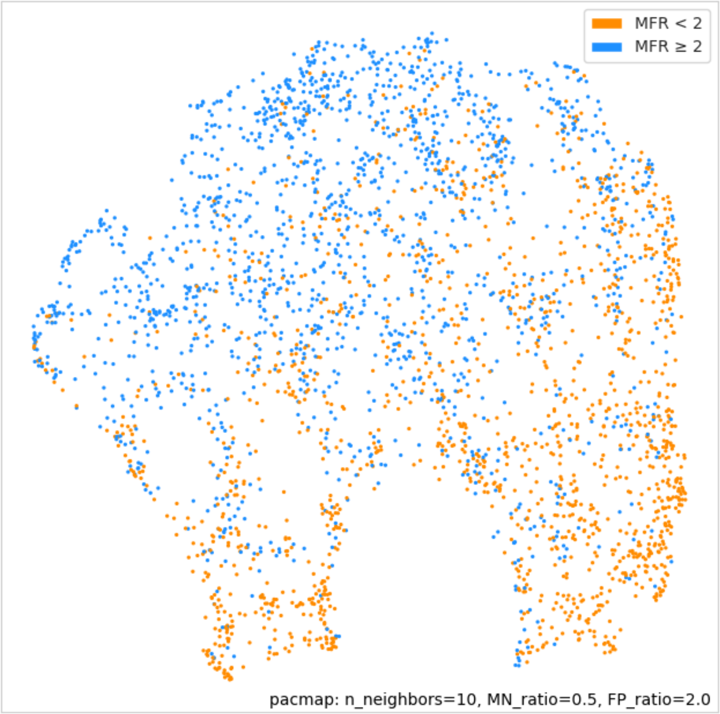
MFR < 2
MFR ≥ 2
pacmap: n_neighbors=10, MN_ratio=0.5, FP_ratio=2.0

*Supplementary Figure S16: 2D projections of feature maps after fine-tuning for the LVEF task on the UM-PET training split. (a) Feature map of stress-rest ECG input to the SSL-pretrained dual 1dViT; (b) feature map of rest-only ECG input to the SSL-pretrained 1dViT; (c) feature map of stress-rest ECG input to the de novo 1dViT (no SSL). Label separation was generally further enhanced compared to that in Supplementary Figure S14b.*

*(a)*

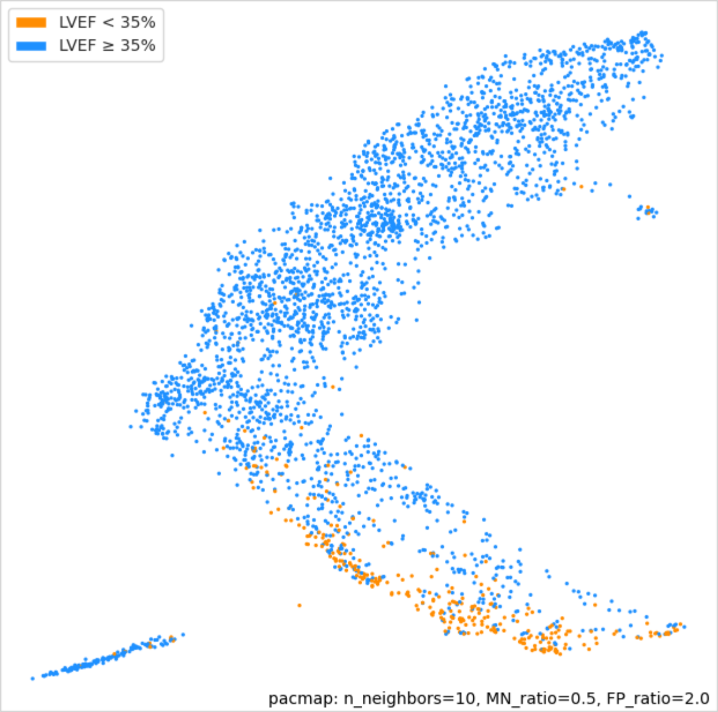


*(b)*

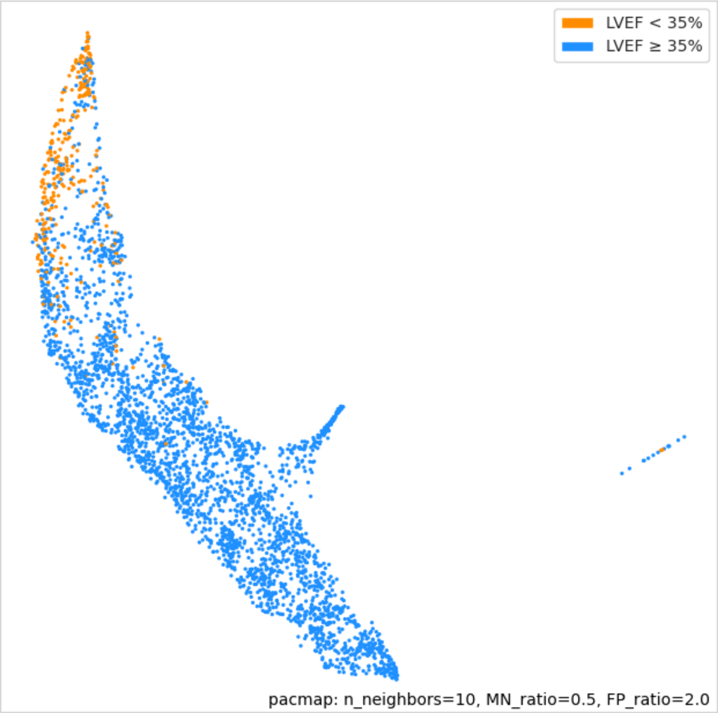

*(c)*

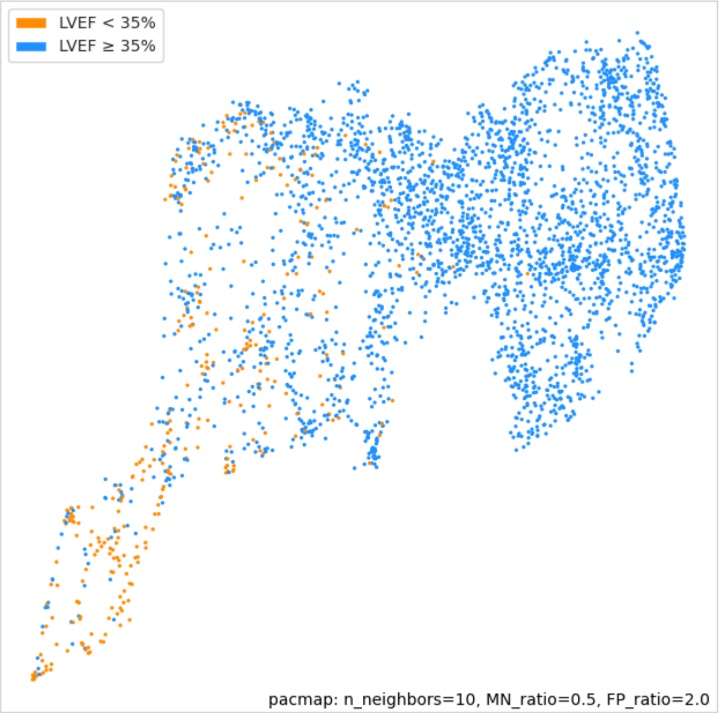

*Supplementary Figure S17: 2D projections of feature maps of stress-rest ECG input to the SSL-pretrained dual 1dViT after fine-tuning for the LVEF task. (a) UM-phSPECT database and (b) UM-exSPECT database. The global cluster structure and pattern of label separation differs compared to UM-PET (Supplementary Figure S16a) due to increasing distribution shift of the SPECT data relative to UM-PET.*

*(a)*

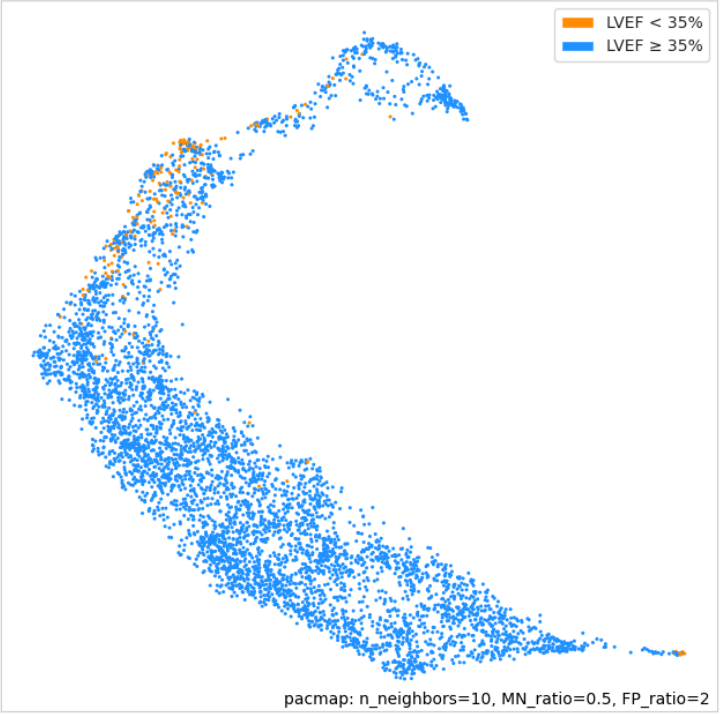


*(b)*

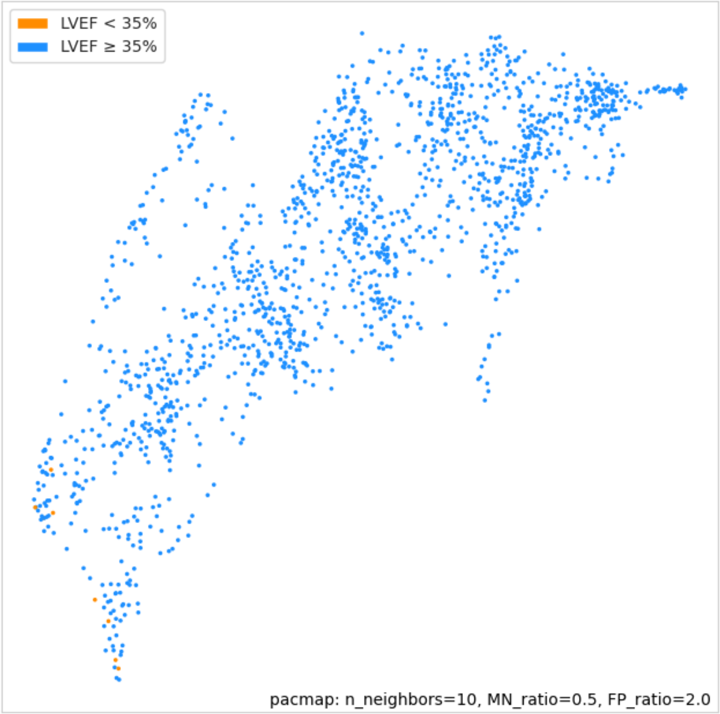
